# Genetic Architecture of 11 Major Psychiatric Disorders at Biobehavioral, Functional Genomic, and Molecular Genetic Levels of Analysis

**DOI:** 10.1101/2020.09.22.20196089

**Authors:** Andrew D. Grotzinger, Travis T. Mallard, Wonuola A. Akingbuwa, Hill F. Ip, Mark J. Adams, Cathryn M. Lewis, Andrew M. McIntosh, Jakob Grove, Søren Dalsgaard, Klaus-Peter Lesch, Nora Strom, Sandra M. Meier, Manuel Mattheisen, Anders D. Børglum, Ole Mors, Gerome Breen, iPSYCH, Tourette Syndrome and Obsessive Compulsive Disorder Working Group of the Psychiatric Genetics Consortium, Bipolar Disorder Working Group of the Psychiatric Genetics Consortium, Major Depressive Disorder Working Group of the Psychiatric Genetics Consortium, Schizophrenia Working Group of the Psychiatric Genetics Consortium, Phil H. Lee, Kenneth S. Kendler, Jordan W. Smoller, Elliot M. Tucker-Drob, Michel G. Nivard

**Author notes:** Correspondence to Andrew D. Grotzinger. These authors jointly directed this work (Elliot M. Tucker-Drob, Michel G. Nivard).

## Abstract

We systematically interrogate the joint genetic architecture of 11 major psychiatric disorders at biobehavioral, functional genomic, and molecular genetic levels of analysis. We identify four broad factors (Neurodevelopmental, Compulsive, Psychotic, and Internalizing) that underlie genetic correlations among the disorders, and test whether these factors adequately explain their genetic correlations with biobehavioral traits. We introduce Stratified Genomic Structural Equation Modelling, which we use to identify gene sets and genomic regions that disproportionately contribute to pleiotropy, including protein-truncating variant intolerant genes expressed in excitatory and GABAergic brain cells that are enriched for pleiotropy between disorders with psychotic features. Multivariate association analyses detect a total of 152 (20 novel) independent loci which act on the four factors, and identify nine loci that act heterogeneously across disorders within a factor. Despite moderate to high genetic correlations across all 11 disorders, we find very little utility of, or evidence for, a single dimension of genetic risk across psychiatric disorders.

---

Psychiatric disorders aggregate both within individuals *and* families. Offspring of parents with psychiatric illness are at higher risk for developing a broad range of psychiatric disorders, not just the specific parental disorder.^1–3^ Moreover, approximately half of individuals with a psychiatric illness will concurrently meet criteria for a second disorder,^4^ and around 40% of individuals will meet diagnostic criteria for four or more psychiatric disorders in their lifetime.^5^ Comorbidity is the norm, rather than the exception. Factor analyses that have modeled these comorbidity patterns consistently identify a transdiagnostic *p*-factor representing general risk across psychiatric disorders, along with several intermediate factors representing more specific clusters of psychiatric risk (*e*.*g*., psychotic disorders, mood disorders).^6–8^ Modern genomics has built on these findings to begin to elucidate the genetic basis for shared risk across disorders,^9,10^ with new statistical tools paired with genome-wide association study (GWAS) data being used to identify pleiotropic variants across disorders.^11,12^ Most recently, Lee *et al*. (2019)^13^ identified three major dimensions of genetic risk sharing (Neurodevelopmental, Compulsive and Psychotic) across eight psychiatric disorders, raising the possibility that key mechanisms of individual disorder risk may operate through these more general factors. Importantly, however, neither phenotypic comorbidity nor genetic correlations among disorders are by themselves sufficient for establishing the etiological, diagnostic, or therapeutic utility of the identified factors.

Here, we apply Genomic Structural Equation Modelling (Genomic SEM) to GWAS data (average total sample size per disorder = 156,771 participants; range = 9,725 - 802,939), to examine the genetic architecture of eleven major psychiatric disorders, across biobehavioral, functional genomic, and molecular genetic levels of analysis. Genomic SEM is able to investigate the multivariate genetic architecture across disorders that could not be measured in the same sample, thereby offering novel insights across the diagnostic spectrum. We begin by estimating several potential genomic factor models, and identify four broad factors that index shared genetic liability within and across disorders. We then evaluate the utility of these factors using a multi-step approach. First, we test the extent to which the factors adequately explain the patterns of genetic correlation between psychiatric disorders and a wide range of external biobehavioral traits specifically selected to represent processes disrupted in psychiatric illness, such as socioeconomic outcomes and cognition. Second, we introduce Stratified Genomic SEM, which we apply to identify gene sets and categories (*e*.*g*., protein-truncating variant-intolerant genes, low MAF SNPs) for which genetic sharing among the disorders, as indexed by each of the factors, is enriched. Finally, we capitalize on Genomic SEM for multivariate GWAS to identify loci that confer risk to multiple disorders via the factors, along with loci that operate heterogeneously across disorders within a given factor. As we observe particularly heterogeneous effects of loci related to problematic alcohol use, we estimate Mendelian randomization models in which pleiotropy is explained by both four latent factors and direct causal influences of problematic alcohol use liability on liability for other psychiatric disorders. Collectively, these results offer critical insights into the shared and disorder-specific mechanisms of genetic risk for psychiatric disease.

## Results

### Factor Analysis of Genetic Covariance across 11 Psychiatric Traits

We curated the most recent European ancestry GWAS summary data for eleven major psychiatric disorders: attention-deficit/hyperactivity disorder (ADHD),^14^ problematic alcohol use (ALCH),^15^ anorexia nervosa (AN),^16^ autism spectrum disorder (AUT),^17^ anxiety disorders (ANX),^18,19^ bipolar disorder (BIP),^20^ major depressive disorder (MDD),^21,22^ obsessive compulsive disorder (OCD),^23^ post-traumatic stress disorder (PTSD),^24,25^ schizophrenia (SCZ), ^26^ and Tourette syndrome (TS).^27^ Data were derived from a range of sources, including the Psychiatric Genomics Consortium (PGC), UK Biobank (UKB), 23andMe, Inc., and iPSYCH (Table S1).

A heatmap of genetic correlations estimated using LD Score regression (LDSC)^9^ across the 11 traits indicates pervasive overlap across disorders, with more pronounced clustering observed among certain constellations of disorders (Figure 1a; Table S2 for LDSC results). We formally modeled this LDSC correlation structure using Genomic SEM by first estimating a series of exploratory factor analyses (EFAs), where the disorders freely load on 2,3,4, or 5 factors, in odd numbered autosomes only. We subsequently fit a series of confirmatory factor analyses (CFAs) specified on the basis of these EFAs, for which model fits were compared using even autosomes only (**Method**). Using odd and even autosome covariance matrices for the EFAs and CFAs, respectively, provided a form of cross-validation to guard against model overfitting.

**Figure 1.**
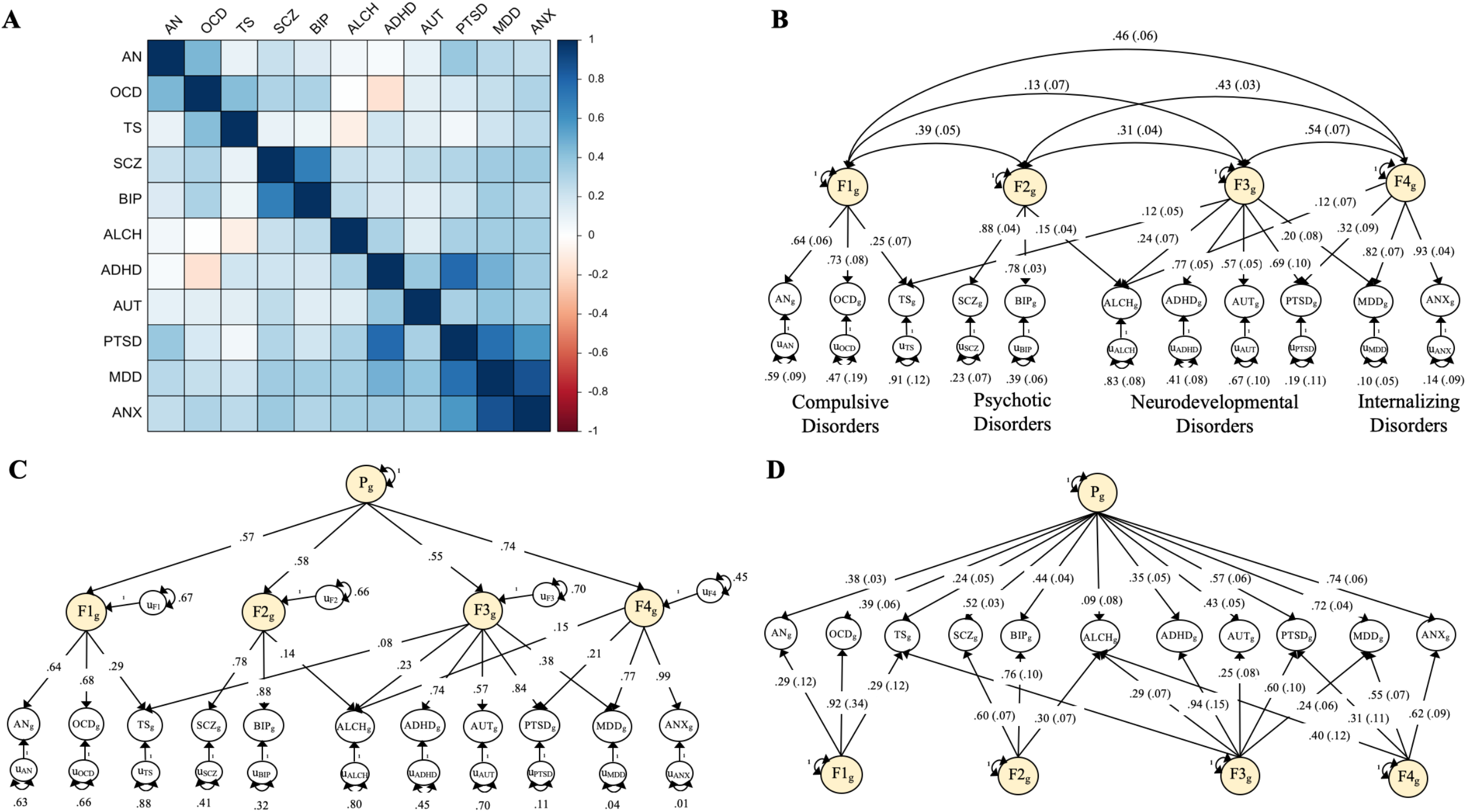
Multivariate Genetic Architecture of 11 Psychiatric Disorders. **Panel A:** Genetic correlations estimated using LDSC. **Panel B:** Standardized results for the correlated factors. **Panel C:** Standardized results from the hierarchical factor model. **Panel D:** Standardized results from the bifactor model. The genetic components of disorders and common genetic factors of disorders are inferred variables that are represented as circles. Regression relationships between variables are depicted as one-headed arrows pointing from the independent variables to the dependent variables. Covariance relationships between variables are represented as two-headed arrows linking the variables. (Residual) variances of a variable are represented as a two-heaed arrow connecting the variable to itself; for simplicity residuals of the indicators are not depicted for the bifactor model. ADHD = attention-deficit/hyperactivity disorder; OCD = obsessive-compulsive disorder; TS = Tourette syndrome; PTSD = post-traumatic stress disorder; AN = anorexia nervosa; AUT = autism spectrum disorder; ALCH = problematic alcohol use; ANX = anxiety; MDD = major depressive disorder; BIP = bipolar disorder; SCZ = schizophrenia.

The best fitting CFA model (for even autosomes: *χ*^2^[33] = 126.85, AIC = 192.85, CFI = .955, SRMR = .078; Table S47 for fit statistics of all models) consisted of four correlated factors (Figure 1b) and, importantly, also fit the data well when fit using all autosomes (*χ*^2^[33] = 161.66, AIC = 227.66, CFI = .975, SRMR = .072). Factor 1 consists of disorders characterized largely by compulsive behaviors (AN, OCD, TS). Factor 2 is characterized by disorders that may have psychotic features (SCZ, BIP). Factor 3 is characterized primarily by childhood-onset neurodevelopmental disorders (ADHD, AUT), but might also be conceptualized as a sensory processing/hyperarousal factor to the extent that PTSD also loads strongly on this factor. Factor 4 is characterized by internalizing disorders (ANX, MDD). These results, with additional disorders and larger GWAS sample sizes, largely replicate findings from PGC Cross-Disorder Group 2 (PGC-CDG2).^13^ More specifically, PGC-CDG2 reported factors representing compulsive, psychotic, and neurodevelopmental disorders, which correspond closely to our first three factors. Our identification of an Internalizing factor can largely be attributed to the inclusion of ANX, and to a lesser extent PTSD, in addition to MDD in the current analysis. It is of note that both TS and ALCH evinced the lowest factor loadings, indicating the most distinct genetic etiology among the 11 disorders in this model.

Cai *et al*. (2020)^28^ have reported that psychiatric phenotypes derived using minimal phenotyping (defined as “individuals’ self-reported symptoms, help seeking, diagnoses or medication”) may produce GWAS signals of low specificity. We therefore conducted a sensitivity analysis in which we excluded GWAS summary statistics for MDD, ANX, ADHD and ALCH that included cohorts with self-report diagnoses or symptoms and refit the correlated factor model. This produced highly similar parameter estimates to those obtained when using all cohorts (Supplementary Results; Figure S1).

The moderate, positive factor intercorrelations observed in Figure 1, in combination with a prior literature indicating a high-order transdiagnostic “*p*-factor”, suggest that a hierarchical factor structure with a single, high-order factor is plausible. Indeed, such a model fit the data well (Figure 1c; even autosomes: *χ*^2^[35] = 173.45, AIC = 235.45, CFI = .933, SRMR = .091; all autosomes: *χ*^2^[35] = 171.37, AIC = 233.37, CFI = .974, SRMR = .079). In this model the *p*- factor explained the greatest proportion of variance in the Internalizing disorders factor (55%) and relatively similar proportions of variance in the remaining three factors (30%-34%). We retain these two key models—the four correlated factors model and the hierarchical factor model—to examine the remaining research questions using data from all autosomes.

### Genetic Correlates of Psychiatric Genetic Factors with External Biobehavioral Traits

A factor model implies a specific causal model, where the factors identified are thought to causally influence their indicators, in this case the 11 psychiatric disorders. Therefore, identified factor structures also imply a certain genetic relationship between external traits and the individual disorders. The degree to which the observed genetic correlation between traits and the psychiatric disorders respect the relationships implied by the factors can be viewed as a validation, or rejection, of the factor structure at one level of analysis. To this end, we examined patterns of correlations across the psychiatric factors and 49 biobehavioral traits relevant to socioeconomic status, anthropomorphic indices, personality, cognitive outcomes, health and disease, risky behavior, and neuropsychiatric outcomes,^29^ 101 metrics of brain morphology,^30^ and circadian activity across 24 hours,^31^ for a total of 174 external traits. Results for brain morphology are presented in the Online Supplement (Figures S3-S4; Table S3), as none of these associations were significant at a Bonferroni corrected threshold for 174 tests (*p* < 2.87E-4).

To evaluate the extent to which each of the 49 biobehavioral traits operated through the factor, we calculated *χ*^2^ difference tests comparing a model in which the trait predicted the factor only, to one in which it predicted the individual disorders of a given factor (or, the first-order factors, in the case of analyses using the *p*-factor model; Figure 2; Figure S5). We term the *χ*^2^ difference across these two models the Q_trait_ heterogeneity index, where a significant index indicates that the pattern of associations between the individual disorders and the external trait is not well-accounted for by the factor. Using a Bonferroni correction, 7/49 correlations were significant for Q_trait_ for the Compulsive factor, 18/49 for the Psychotic factor, 39/49 for the Neurodevelopmental factor, 17/49 for the Internalizing factor, and 38/49 for the *p*-factor (Table S4). Excluding significant Q_trait_ correlations (i.e., correlations not operating through the factor), and using the same Bonferroni correction, 17 correlations were significant for the Compulsive factor, 12 for the Psychotic factor, 5 for the Neurodevelopmental factor, 20 for the Internalizing factor, and 3 for the *p*-factor.

**Figure 2.**
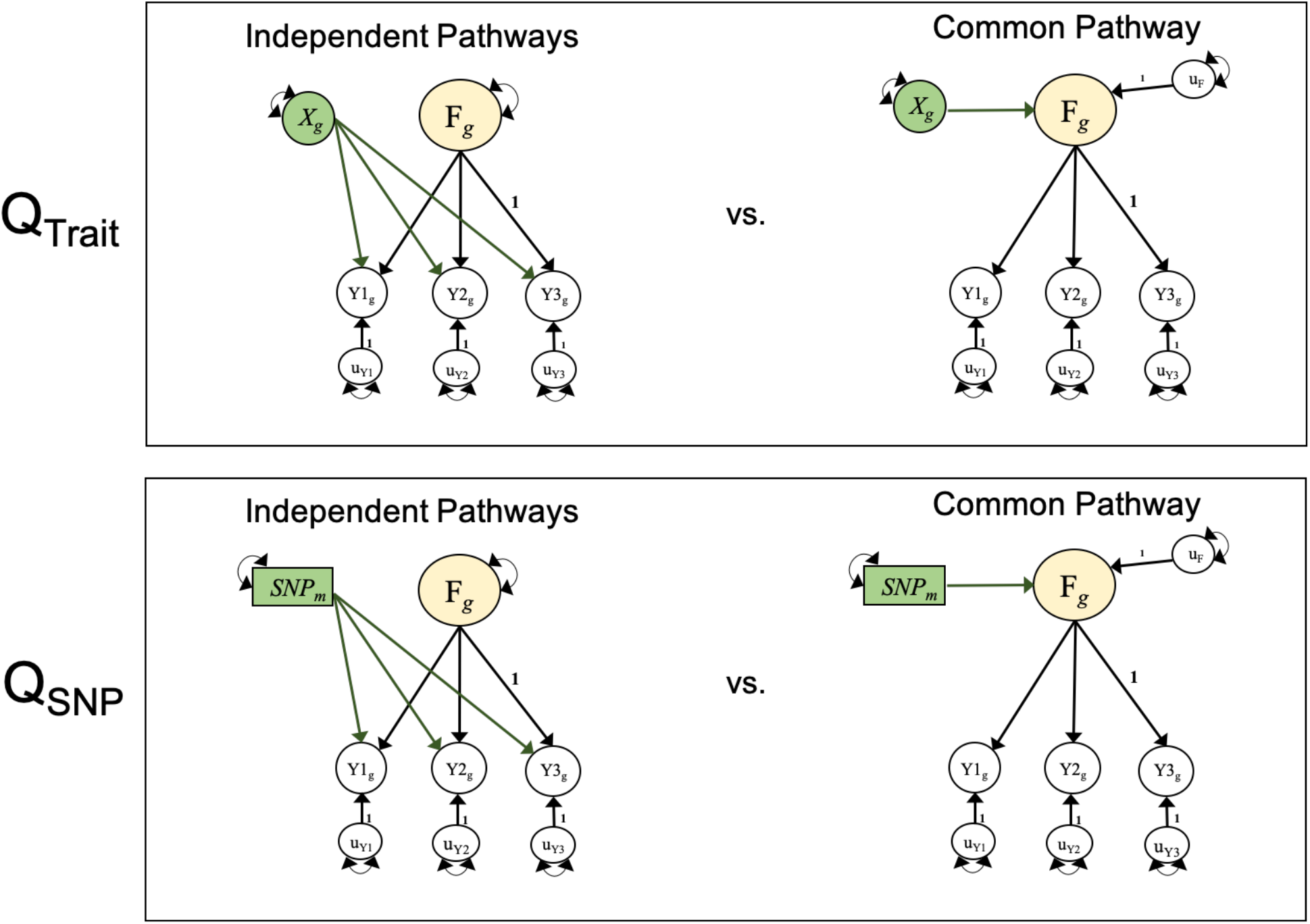
Model Comparisons for Producing Q metrics. Unstandardized path diagrams for *common pathway* (left) and *independent pathways* (right) models used to compute the Genomic SEM heterogeneity statistics for associations with external traits (Q_Trait_, top) and individual SNPs (Q_SNP_, bottom). In this example, F is a common genetic factor of the genetic components of 3 GWAS phenotypes (Y_1_-Y_3_). Observed variables are represented as squares and latent variables are represented as circles. The genetic component of each phenotype is represented with a circle as the genetic component is a latent variable that is not directly measured, but is inferred using LDSC. SNPs are directly measured, and are therefore represented as squares. Single-headed arrows are regression relations, and double-headed arrows are variances. Paths labeled 1 are fixed to 1 for model identification purposes. All unlabeled paths represent freely estimated model parameters. Q represents the decrement in model fit of the *common pathway* model relative to the more restrictive *independent pathways* model. Q is a χ^2^ distributed test statistic with k-1 degrees of freedom, representing the difference between the *k* SNP-phenotype or Trait-phenotype *b* coefficients in the independent pathways model and the 1 SNP-factor or Trait-factor *b* coefficient in the *common pathway* model. Q_Trait_ indexes whether the pattern of genetic associations between the genetic component of an external trait (depicted as X_g_) and the individual disorders is well accounted for by a given factor. Q_SNP,_ indexes whether the associations between an individaul SNP (depicted as SNP_m_) and the individual dissorders is well accounted for by the factor. For simplicity, we depict a stylized representation containing only one factor and three disorders. The full models used to derive Q_Trait_ and Q_SNP_ for the empirical analyses reported in this paper are presented in Figures S5 and S38.

As expected, all factors were positively genetically associated with psychiatric phenotypes from outside studies, including the cross-disorder iPSYCH results, and negatively genetically correlated with indices of positive mental health (*e*.*g*., subjective well-being, family relationship satisfaction; Figure S6). In the remainder of this section, we generally describe patterns of genetic correlations with external biobehavioral traits outside of the psychiatric domain.

The Compulsive disorders factor was negatively genetically correlated with anthropomorphic traits (BMI, waist-to-hip ratio) and risk-taking behaviors (e.g., automobile speeding, pub attendance; Figure 3). Educational attainment (EA) evinced a particular pattern of genetic associations with the individual compulsive disorders that were inconsistent with their operation via the Compulsive disorders factor, where AN was more positively associated relative to OCD and TS (Figure S7).

**Figure 3.**
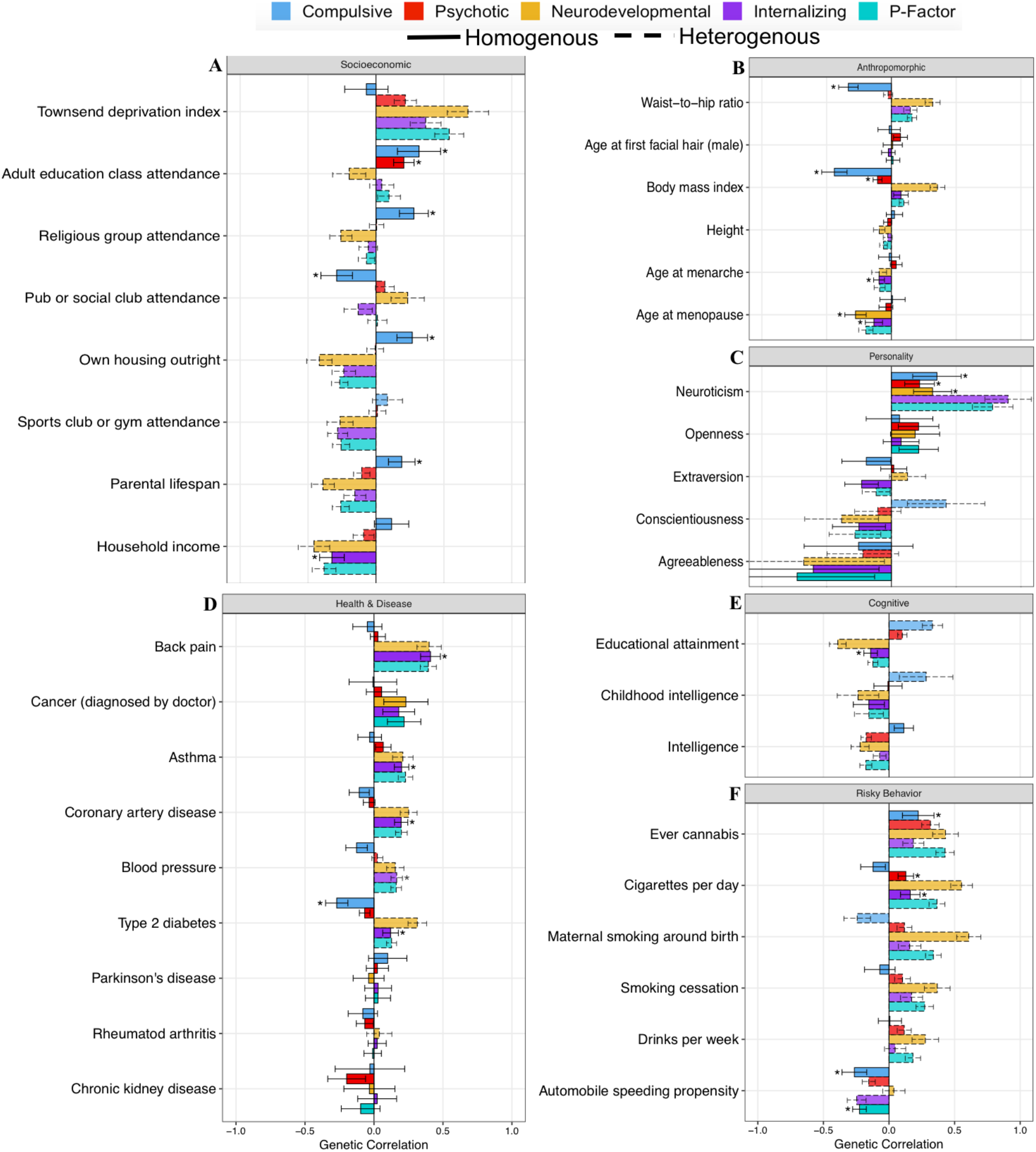
Genetic Correlations with Complex Traits across Psychiatric Factors. Panels depict genetic correlations with complex traits of interest for the four psychiatric factors from the correlated factors model and the second-order, *p*-factor from the hierarchical model. Genetic correlations are shown for socioeconomic (**Panel A**), anthropromorphic (**Panel B**), personality (**Panel C**), health and disease (**Panel D**), cognitive (**Panel E**), and risky behavior outcomes (**Panel F**). Bars depicted with a dashed outline were significant at a Bonferroni corrected threshold for model comparisons indicating heterogeneity across the factor indicators in their genetic correlations with the outside trait. Error bars reflect 95% confidence intervals. Bars depicted with an * above produced a genetic correlation that was significant at a Bonferroni corrected threshold and were not significantly heterogeneous.

The Psychotic disorders factor was negatively associated with obesity related outcomes (BMI, Type 2 diabetes) and positively associated with neuroticism. Phenotypes whose patterns of genetic associations with the individual disorders were inconsistent with their operation via the Psychotic disorders factor were substance use phenotypes (*e*.*g*., drinks per week, cannabis use), for which genetic associations with SCZ were more pronounced than with BIP, and cognitive (*e*.*g*., EA) and risk-taking phenotypes (*e*.*g*., automobile speeding), for which BIP exhibited more pronounced positive associations.

The Neurodevelopmental disorders factor was genetically associated with earlier age at menopause. All other external correlates outside of the psychiatric domain that survived Bonferroni-correction exhibited patterns of associations with the individual neurodevelopmental disorders that were inconsistent with their operation via the factor. Cognitive (*e*.*g*., educational attainment, intelligence), anthropometric (e.g., BMI), and economic outcomes (*e*.*g*., Townsend deprivation) had the strongest disorder-specific associations, with positive associations observed for AUT, and negative associations for PTSD and ADHD. The Neurodevelopmental disorders factor therefore performed poorly at this level of validation due largely to divergent patterns for AUT.

The Internalizing disorders factor exhibited negative genetic associations with extraversion, age at menopause, EA, and positive associations with various adverse health outcomes (e.g., asthma, back pain, coronary artery disease). Phenotypes with the strongest disorder-specific associations included socioeconomic phenotypes (*e*.*g*., owning a house outright), which tended to exhibit more pronounced negative genetic associations with MDD than with ANX.

The *p*-factor exhibited a homogenous genetic correlation with automobile speeding propensity. All other external non-psychiatric correlates that survived Bonferroni-correction exhibited patterns of associations with the first order psychiatric genetic factors that were inconsistent with their operation via the *p*-factor. The genetic associations with EA deviated most strongly from the hierarchical factor structure. These patterns of widespread heterogeneity in genetic correlations with external phenotypes undermine the utility of the *p*-factor.

#### Accelerometer Data

Atypical patterns of physical movement throughout the 24-hour cycle may reflect disturbances in basic homeostatic processes that confer transdiagnostic psychiatric risk.^32^ Using accelerometer data from UKB,^31^ we next examined genetic correlations between the individual psychiatric traits and factors and physical movement across a 24-hour period (Figure 4; Table S5). The same Q_trait_ indices described for complex traits were used to determine whether patterns of associations with hours of movement were well-accounted for by the factors. Using a Bonferroni correction for 174 tests, 1 correlation was significant for Q_trait_ for the Compulsive factor, 2 for the Psychotic factor, 12 for the Neurodevelopmental factor, 7 for the Internalizing factor, and 18 for the *p*-factor. Excluding any significant Q_trait_ correlations, and using the same Bonferroni correction, 8 correlations were significant for the Compulsive factor, 4 for the Psychotic factor, 1 for the Neurodevelopmental factor, 6 for the Internalizing factor, and 2 for the *p*-factor.

**Figure 4.**
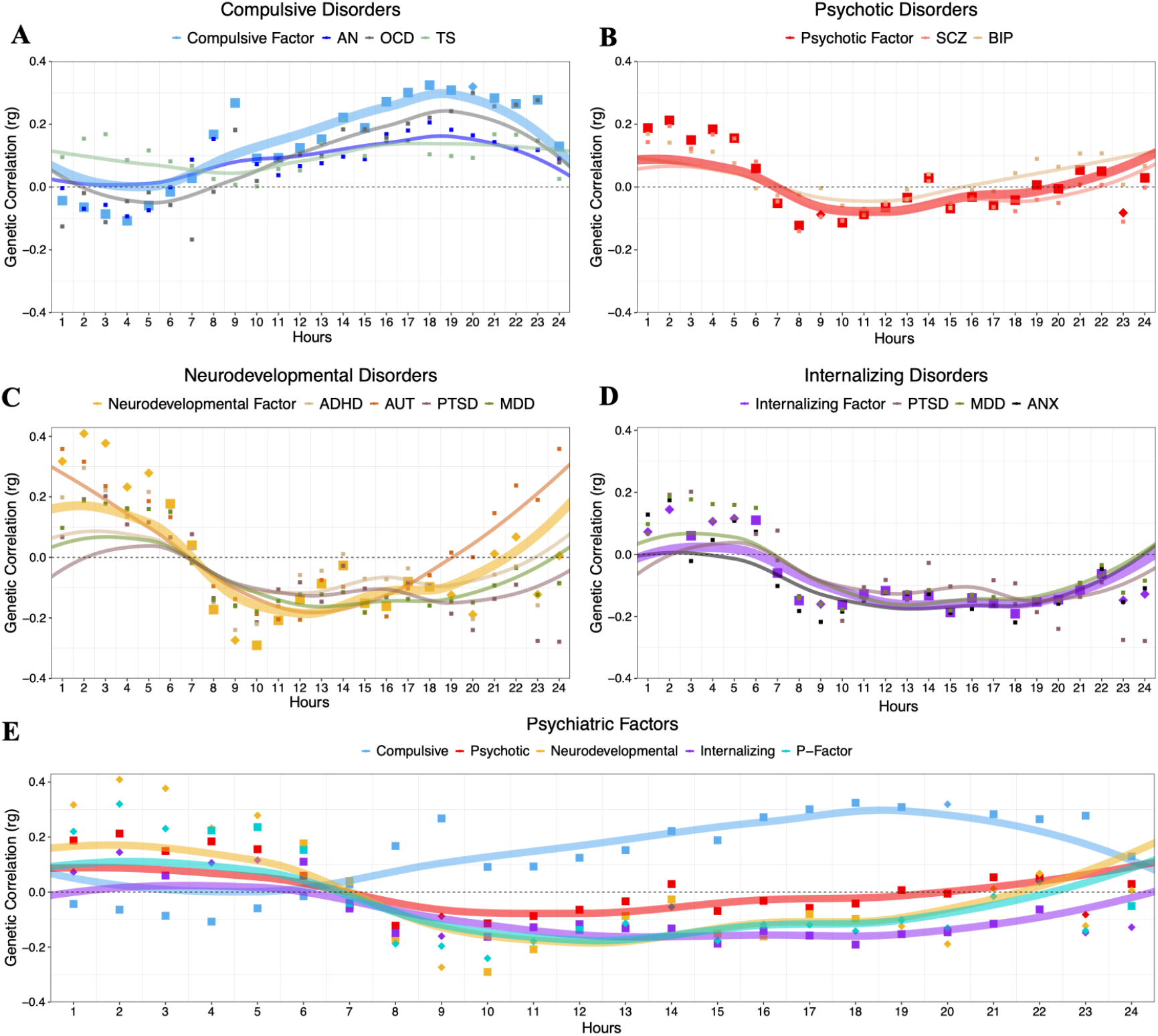
Genetic Correlations with Accelerometer Data across Psychiatric Disorders and Factors. Panels depicts genetic correlations between accelerometer-based average total hourly movement within the 24-hr day beginning at midnight (*N*∼95,000) and each psychiatric disorder, along with the respective psychiatric factor, for the compulsive disorders (**Panel A**), psychotic disorders (**Panel B**), neurodevelopmental disorders (**Panel C**), internalizing disorders (**Panel D**), and psychiatric factors (**Panel E**). Across all panels, the psychiatric factors are depicted with larger points and lines. For the psychiatric factors, points depicted as diamonds were significant at a Bonferroni corrected threshold for model comparisons indicating heterogeneity across the factor indicators in their genetic correlations with that particular time point. As it loaded on three different factors (cf. Figure 1), ALCH was not as assigned to a panel above. Lines represent loess regression lines estimated in *ggplot2*.

Compulsive disorders were positively genetically correlated with physical movement throughout the daylight hours and into the evening. Psychotic disorders were positively genetically correlated with excess movement in the early morning hours. The pattern of associations deviated from the factor structure largely in the daylight and evening hours, with larger positive genetic correlations observed for BIP. Genetic correlations with movement throughout the day where heterogenous across disorders that load on the Neurodevelopmental disorders factor. This was primarily due to unique associations for AUT, for which positive genetic correlations were observed during the evening hours relative to negative correlations for other disorders. Internalizing disorders were negatively genetically correlated with movement throughout the daylight and earlier evening hours.

### Genetic Enrichment of Psychiatric Genetic Factors (Stratified Genomic SEM)

We developed Stratified Genomic SEM to allow the basic principles of Genomic SEM to be applied to genetic covariance matrices estimated in different gene sets and categories (**Method**). These gene sets and categories, collectively referred to as annotations, can be constructed based on a variety of sources, such as collateral gene expression data obtained from single-cell RNA sequencing. Such an analysis goes beyond methods such as Stratified LDSC^33^ that estimate enrichment of heritability for particular traits within functional annotations. Rather, Stratified Genomic SEM allows us to ask whether pleiotropic loci are enriched within particular annotations.

We fit Stratified Genomic SEM models that allowed variances of the common genetic factors, and disorder-specific effects, to vary across annotations to examine whether the degree of risk sharing and differentiation is enriched across disorders. Enrichment is defined as the ratio of the proportion of genome-wide risk sharing indexed by the annotation to that annotation’s size as a proportion of the genome (**Method**). The null, corresponding to no enrichment, is a ratio of 1.0, with values above 1.0 indicating enrichment of pleiotropic signal within a functional annotation. We included functional annotations from the most recent 1000 Genomes Phase 3 BaselineLD Version 2.2,^34^ for tissue specific histone marks based on data from the Roadmap Epigenetics Project,^35^ for specific gene expression constructed based on RNA sequencing data from human tissues from GTEx,^36^ and for annotations constructed from human, mouse, and rat microarray experiments (*i*.*e*., DEPICT).^37^ In addition, we created 29 annotations to examine the interaction between expression patterns for protein-truncating variant (PTV)-intolerant (PI) genes (obtained from the Genome Aggregation Database; gnomAD^38^), and human brain cells in the hippocampus and prefrontal cortex (obtained from GTEx^39^). In total, enrichment analyses were based on 168 binary annotations. Using a Bonferroni correction for 168 tests, we identify 40 annotations that were significantly enriched for the Psychotic disorders factor, 1 annotation (conserved primate) for the Neurodevelopmental disorders factor, 4 annotations for the Internalizing disorders factor, and 38 annotations for the *p*-factor (Table S6).

PI results revealed that these annotations were particularly enriched for the Psychotic disorders factor, with 5 out of the 10 most significantly enriched gene sets falling in this category (Figure 5). Moreover, we observe that specific intersections of PI and brain cells were more enriched than others, with the interaction of PI genes and genes expressed for excitatory (*e*.*g*., hippocampal CA1 neurons) and GABAergic neurons displaying the most significant enrichment for the Psychotic disorders factor. PI genes reflect a broad functional class that has been found to confer risk across a wide array of disorders (e.g., AUT, ADHD, BIP and SCZ^40^). These findings thus offer insight into neuronal subcategories within the overarching PI gene set that are specifically associated with shared risk across BIP and SCZ.

**Figure 5.**
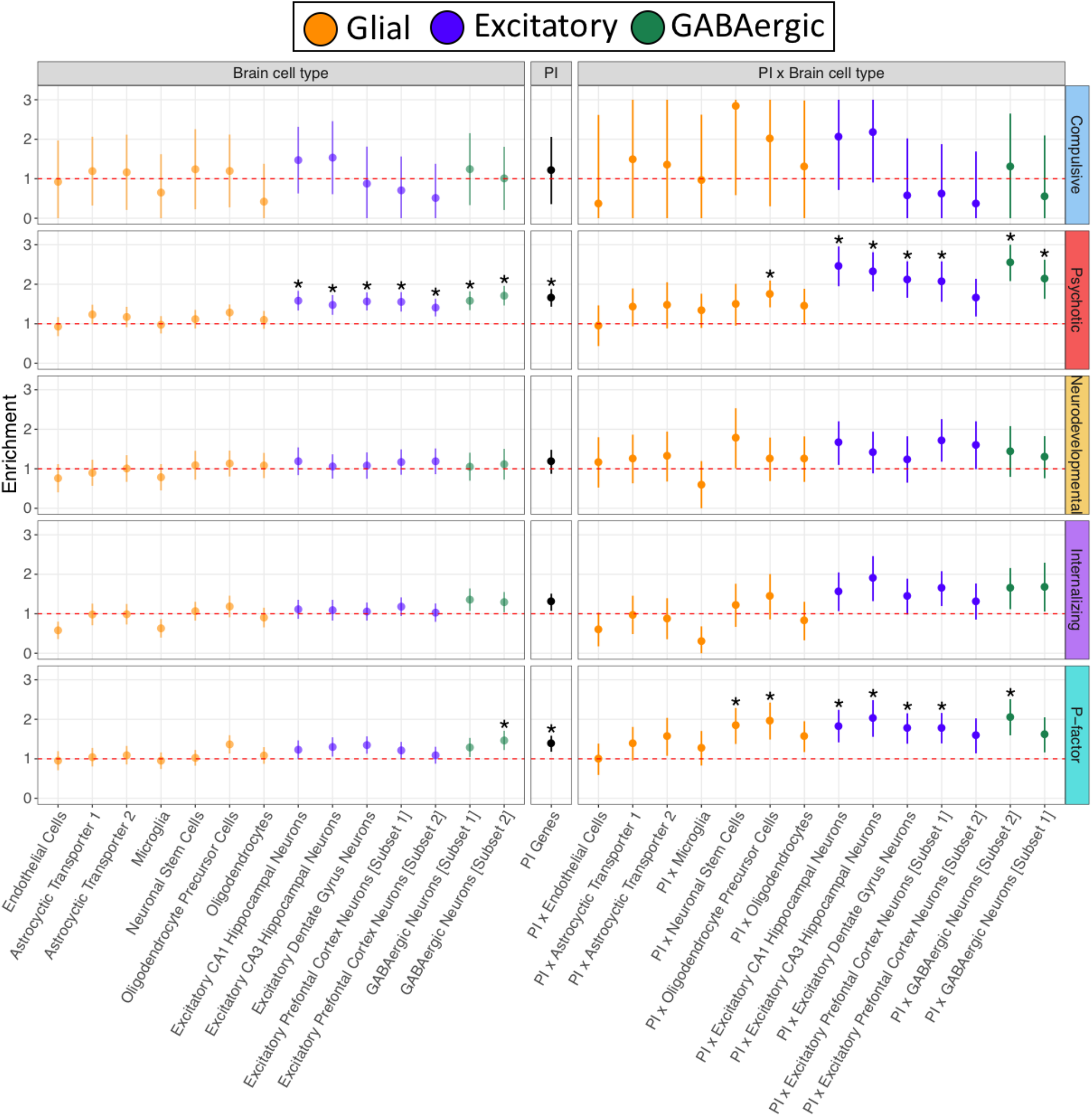
Genetic Enrichment of Factors for Brain Cell, PI, and PI × Brain Cell Annotations. Figure depicts enrichment of the four factors from correlated factors model and the second-order, *p*-factor from the hierarchical factor model for the brain cell genes, protein-truncating variant (PTV)-intolerant (PI) genes, and PI × brain cell gene annotations. Enrichment is indexed by the ratio of the proportion of genome-wide relative risk sharing indexed by the annotation to that annotation’s size as a proportion of the genome. The red dashed line reflects the null ratio of 1.0, corresponding to no enrichment. Ratios greater than 1.0 indicate enrichment of pleiotropic signal whereas ratios less than 1.0 indicate depletion of pleiotropic signal. Error bars depict 95% confidence intervals. Points depicted with a * were significantly enriched at a Bonferroni corrected threshold. To maintain equal scaling purposes across all panels, error bars are capped at 3 and 0 for the Compulsive disorders factor; no annotations were significant for this factor.

We find that shared genetic variance across disorders, as estimated by a higher order *p*-factor, is enriched in conserved annotations (e.g., conserved primate; Genomic Evolutionary Rate Profiling [GERP]) and that enrichment increases from low to high MAF alleles (Figure S8-S14). This indicates that previous reports of similar findings for individual disorders^33,41^ may reflect enrichment of pleiotropic variants that are broadly relevant for many disorders. The most enriched annotations for the Neurodevelopmental and Internalizing disorders factors were fetal female brain DNase and fetal male brain H3K4me1, respectively, both of which have been previously reported to be enriched for general liability across psychiatric disorders.^42^ For specific tissues, we observe that brain regions are generally enriched, as is also observed for other complex traits,^43^ but were most enriched for the Psychotic disorders factor.

Results for genetic enrichment of the residuals of the psychiatric factors after accounting for variance explained by the *p*-factor are presented in the **Online Supplement** (Figures S15-S20; Table S5). These results indicated slightly attenuated signal across enrichment categories relative to enrichment from the correlated factors model, with one important exception: the enrichment signal was even stronger in the PI × neuronal annotations when examining the variance in the Psychotic disorders factor that was unique of the three remaining factors. This provides compelling evidence that variants within PI genes expressed in specific hippocampal and prefrontal cortex neuronal cells are distinctly important for genetic overlap between BIP and SCZ.

### Unstructured Multivariate GWAS

We went on to conduct an unstructured multivariate GWAS that computes an omnibus index of association across all 11 disorders. This GWAS was conducted within Genomic SEM by comparing a maximally complex model in which the SNP is allowed to have direct regression relations with each of the 11 disorders against a null model in which the SNP is associated with none of the disorders. This omnibus test is *χ*^2^ distributed with 11 *df*, and quantifies evidence for an overall effect of the SNP on any subset of the disorders, irrespective of the patterning or directionality of the effects. We refer to this as an unstructured multivariate GWAS because the tested model freely estimates as many SNP regressions as there are disorders, and can identify variants associated with a subset of the psychiatric disorders regardless of their loading on the higher order factors we observed.

The unstructured multivariate GWAS identified 184 associated loci, 39 of which were not in LD with any of the univariate associations (Figure 6 for Miami plots; Figure S21 for QQ-plots; Table S7 for individual hits). Of these 39 novel hits, nine have not been described for independent studies of psychiatric traits/symptoms and were largely characterized by hits previously found for cognitive (e.g., intelligence) or anthropometric traits (e.g., BMI; Table S7). Moreover, 7 hits were entirely novel in that they were not in LD with any previously discovered hits in the GWAS catalogue. For comparative purposes, we consider overlap with the 109 pleiotropic and 146 total hits from PGC-CDG2^13^ given both overlapping datasets and research questions. The unstructured multivariate GWAS recaptures 69 of the 109 (63.3%; Table S8) pleotropic loci and 97 of the 146 (66.4%) total loci from PGC-CDG2.

**Figure 6.**
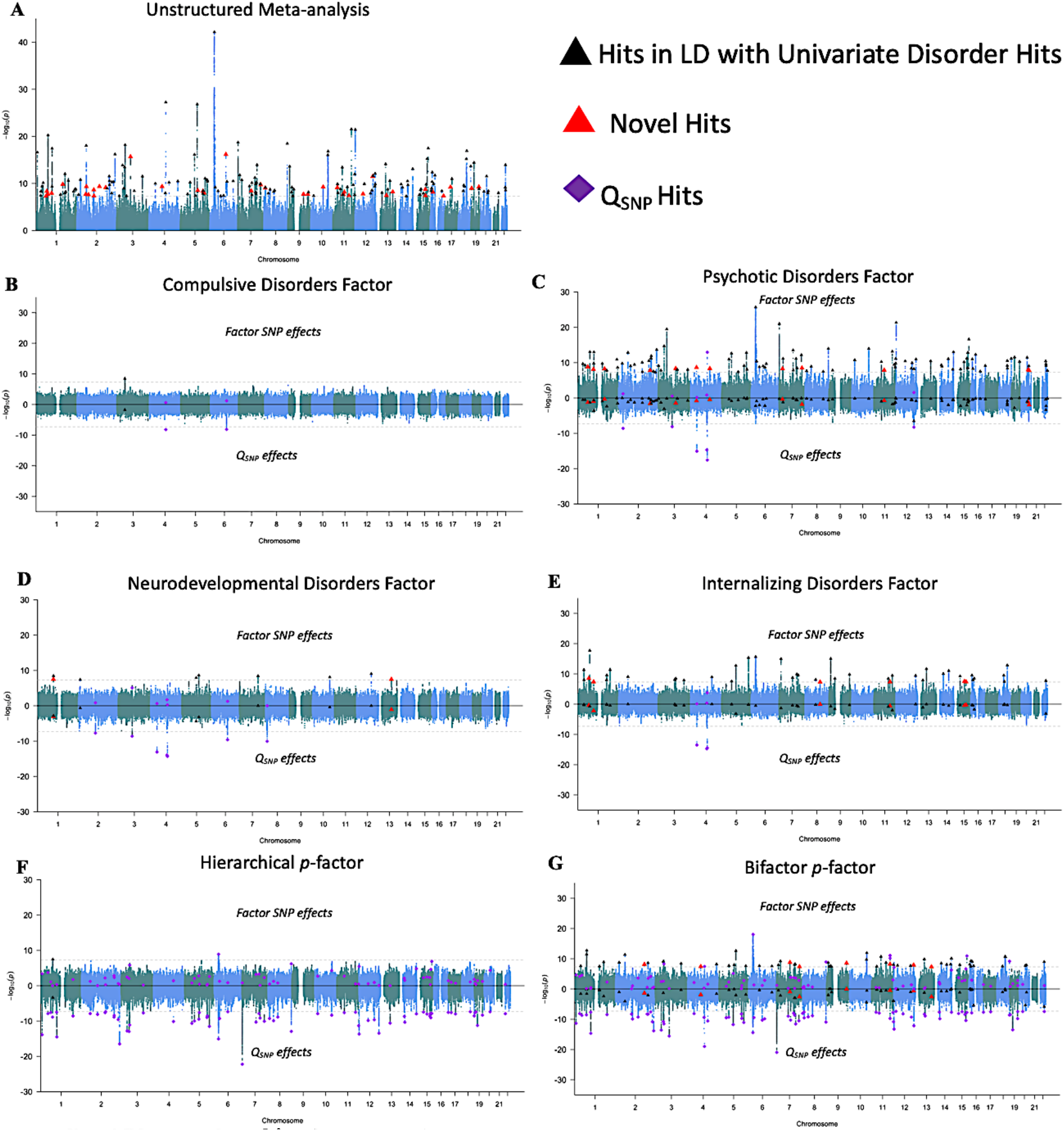
Miami Plots for Psychiatric Factors. **Panel A** depicts results from an unstructured meta-analysis of the 11 psychiatric traits (**Panel A**). Results from the correlated factors model are depicted for the Compulsive disorders factor (Factor 1; **Panel B**), Psychotic disorders factor (Factor 2; **Panel C**), Neurodevelopmental disorders factor (Factor 3; panel D), and Internalizing disorders factor (Factor 4; **Panel E**). **Panel F** depicts the results of the SNP effect on the second-order *p*-factor from the hierarchical model. **Panel G** depicts results from a model in which the SNP predicted the *p*-factor from a bifactor model. The top half of the plots depict the −log10(p) values for SNP effects on the factor; the bottom half depicts the log10(p) values for the factor specific Q_SNP_ effects. As the omnibus meta-analysis does not impose a structure on the patterning of SNP-disorder associations, it does not have a Q_SNP_ statistic. The gray dashed line marks the threshold for genome-wide significance (*p* < 5 × 10^−8^). Black triangles denote independent factor hits that were in LD with hits for one of the univariate indicators and were not in LD with factor-specific Q_SNP_ hits. Large red triangles denote novel loci that were not in LD with any of the univariate GWAS or factor-specific Q_SNP_ hits. Purple diamonds denote Q_SNP_ hits.

### Structured Multivariate GWAS

We used Genomic SEM to perform two structured multivariate GWASs, one using the correlated factors model (with Factors 1-4 as the GWAS target), and one using the hierarchical factor model (with the higher order *p*-factor as the GWAS target; Figure 6). We refer to these multivariate GWASs as structured, because the different models used for each define a specific pattern, or *structure*, of the relationship between the SNP and the 11 disorders. For each of the two multivariate GWASs, and for each factor used as a GWAS target, we estimate SNP-specific indices of heterogeneity with the Q_SNP_^44^ statistic that indexes violation of the null hypothesis that the SNP acts on the individual disorders entirely via the factor on which they load (Figure 3; see **Method**). A Q_SNP_ statistic is typically significant when the SNP effect is highly specific to an individual disorder or when SNP effects are highly heterogeneous across disorders, such as when there is divergent directionality across the disorders. Thus, we use these GWASs to identify whether variants specifically relate to broad-dimensions of genetic risk, or to a specific disorder or disorders. Corresponding results from using the genome-wide S-LDSC matrix can be found in Tables S27-S39 and Figures S22-S23. LDSC and S-LDSC produced highly similar multivariate GWAS results (Supplementary Results; Figures S24-25). Polar plots of individual variants estimated as genome-wide significant are presented in Figure S26.

We identified 1 hit for the Compulsive disorders factor, a locus also associated with AN^16^ (Table S9-S10). We identify two loci for the Compulsive disorders factor-specific Q_SNP_ statistic (Table S11), including a locus (rs1906252) with strong opposing effects on AN and TS.

We identified 108 hits for the Psychotic disorders factor, 96 of which were in LD with previously reported associations with BIP^45^ and SCZ (Table S12), and 12 of which were novel relative to the contributing univariate GWASs. Of these 12 unique hits, 8 have been reported as hits in independent (or semi-independent) external GWAS of psychiatric traits, 2 were novel for psychiatric traits, and 2 were entirely novel (Table S13). Psychotic disorders, factor-specific Q_SNP_ statistic revealed 6 hits, 3 of which were in LD with hits for ALCH (Table S14), including a locus in the well-described Alcohol Dehydrogenase 1B (ADH1B) gene that was significant for factor-specific Q_SNP_ for all four factors.

We identified nine hits for the Neurodevelopmental disorders factor (Table S15), 3 of which were in LD with hits for ADHD^14^ or MDD, and 2 of which were novel relative to the contributing univariate GWASs. These two novel hits were in LD with hits previously described for GWAS of psychiatric traits (Table S16). There were 7 hits for the Neurodevelopmental Q_SNP_ statistic, many of which appeared to be specific to AUT^17^ (Table S17).

We identified 44 independent hits for the Internalizing disorders factor, 6 of which were unique of hits from the contributing univariate GWASs (Table S18). Among these 6 novel loci, 3 were identified in outside studies of psychiatric traits, one has been identified for smoking initiation, and two have yet to be described for any trait (Table S19). Three loci were identified for the Internalizing factor-specific Q_SNP_ statistic, all three of which were in LD with hits for ALCH (Table S20). We note that the discrepancy in the number of univariate MDD hits (109) relative to the number of Internalizing factor hits (44) can be attributed to a combination of signal specific to MDD and splitting the MDD signal across two factors (Figure S27).

Of the 109 pleiotropic hits from PGC-CDG2, none were in LD with hits for the Compulsive disorders factors, 52 hits were in LD with hits for the Psychotic disorders factor, 4 hits were in LD with hits for the Neurodevelopmental disorders factor, and 14 hits were in LD with hits for the Internalizing disorders factor. As 5 of these overlapping hits were redundant across the factors, the correlated factors model indicates that 65 of the 109 (59.6%) PGC-CDG2 hits may be interpreted as acting pleiotropically via the factors identified here. Nine hits from the correlated factors model were in LD across the factors, and 1 hit was in LD with a Q_SNP_ hit. In total, we therefore discover 152 independent loci that are likely to operate through pleiotropic mechanisms, 20 of which that were novel relative to the univariate traits. Accounting for LD across factor-specific Q_SNP_ hits, we identify nine independent Q_SNP_ hits that do not conform to the identified factor structure (Table 1), a third of which appeared to operate through pathways unique to ALCH.

**Table 1.**
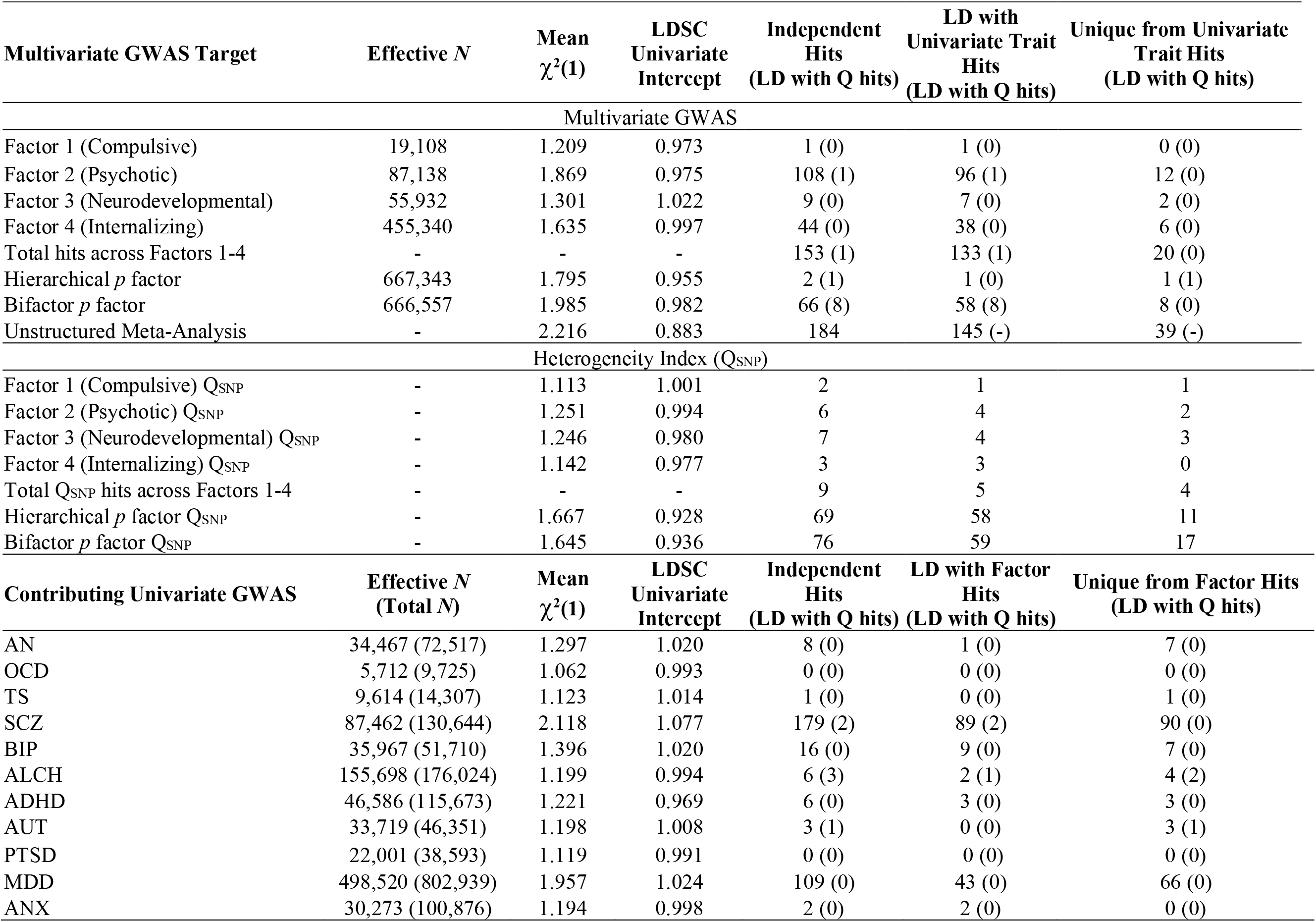
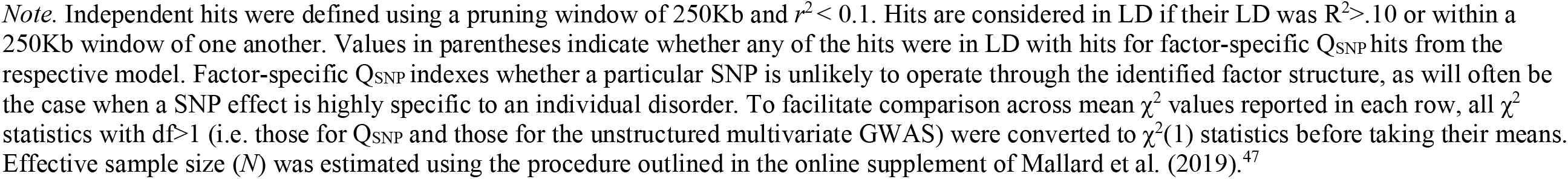
Genome-wide Multivariate GWAS Results

| Multivariate GWAS Target | Effective *N* | Mean $\chi^2(1)$ | LDSC Univariate Intercept | Independent Hits (LD with Q hits) | LD with Univariate Trait Hits (LD with Q hits) | Unique from Univariate Trait Hits (LD with Q hits) |
| --- | --- | --- | --- | --- | --- | --- |
| Multivariate GWAS |  |  |  |  |  |  |
| Factor 1 (Compulsive) | 19,108 | 1.209 | 0.973 | 1 (0) | 1 (0) | 0 (0) |
| Factor 2 (Psychotic) | 87,138 | 1.869 | 0.975 | 108 (1) | 96 (1) | 12 (0) |
| Factor 3 (Neurodevelopmental) | 55,932 | 1.301 | 1.022 | 9 (0) | 7 (0) | 2 (0) |
| Factor 4 (Internalizing) | 455,340 | 1.635 | 0.997 | 44 (0) | 38 (0) | 6 (0) |
| Total hits across Factors 1-4 | - | - | - | 153 (1) | 133 (1) | 20 (0) |
| Hierarchical *p* factor | 667,343 | 1.795 | 0.955 | 2 (1) | 1 (0) | 1 (1) |
| Bifactor *p* factor | 666,557 | 1.985 | 0.982 | 66 (8) | 58 (8) | 8 (0) |
| Unstructured Meta-Analysis | - | 2.216 | 0.883 | 184 | 145 (-) | 39 (-) |
| Heterogeneity Index ($Q_{\text{SNP}}$) | | | | | | |
| Factor 1 (Compulsive) $Q_{\text{SNP}}$ | - | 1.113 | 1.001 | 2 | 1 | 1 |
| Factor 2 (Psychotic) $Q_{\text{SNP}}$ | - | 1.251 | 0.994 | 6 | 4 | 2 |
| Factor 3 (Neurodevelopmental) $Q_{\text{SNP}}$ | - | 1.246 | 0.980 | 7 | 4 | 3 |
| Factor 4 (Internalizing) $Q_{\text{SNP}}$ | - | 1.142 | 0.977 | 3 | 3 | 0 |
| Total $Q_{\text{SNP}}$ hits across Factors 1-4 | - | - | - | 9 | 5 | 4 |
| Hierarchical *p* factor $Q_{\text{SNP}}$ | - | 1.667 | 0.928 | 69 | 58 | 11 |
| Bifactor *p* factor $Q_{\text{SNP}}$ | - | 1.645 | 0.936 | 76 | 59 | 17 |
| Contributing Univariate GWAS | Effective *N* (Total *N*) | Mean $\chi^2(1)$ | LDSC Univariate Intercept | Independent Hits (LD with Q hits) | LD with Factor Hits (LD with Q hits) | Unique from Factor Hits (LD with Q hits) |
| AN | 34,467 (72,517) | 1.297 | 1.020 | 8 (0) | 1 (0) | 7 (0) |
| OCD | 5,712 (9,725) | 1.062 | 0.993 | 0 (0) | 0 (0) | 0 (0) |
| TS | 9,614 (14,307) | 1.123 | 1.014 | 1 (0) | 0 (0) | 1 (0) |
| SCZ | 87,462 (130,644) | 2.118 | 1.077 | 179 (2) | 89 (2) | 90 (0) |
| BIP | 35,967 (51,710) | 1.396 | 1.020 | 16 (0) | 9 (0) | 7 (0) |
| ALCH | 155,698 (176,024) | 1.199 | 0.994 | 6 (3) | 2 (1) | 4 (2) |
| ADHD | 46,586 (115,673) | 1.221 | 0.969 | 6 (0) | 3 (0) | 3 (0) |
| AUT | 33,719 (46,351) | 1.198 | 1.008 | 3 (1) | 0 (0) | 3 (1) |
| PTSD | 22,001 (38,593) | 1.119 | 0.991 | 0 (0) | 0 (0) | 0 (0) |
| MDD | 498,520 (802,939) | 1.957 | 1.024 | 109 (0) | 43 (0) | 66 (0) |
| ANX | 30,273 (100,876) | 1.194 | 0.998 | 2 (0) | 2 (0) | 0 (0) |

*Note.* Independent hits were defined using a pruning window of 250Kb and $r^2 < 0.1$ . Hits are considered in LD if their LD was $R^2 > .10$ or within a 250Kb window of one another. Values in parentheses indicate whether any of the hits were in LD with hits for factor-specific $Q_{\text{SNP}}$ hits from the respective model. Factor-specific $Q_{\text{SNP}}$ indexes whether a particular SNP is unlikely to operate through the identified factor structure, as will often be the case when a SNP effect is highly specific to an individual disorder. To facilitate comparison across mean $\chi^2$ values reported in each row, all $\chi^2$ statistics with $df > 1$ (i.e. those for $Q_{\text{SNP}}$ and those for the unstructured multivariate GWAS) were converted to $\chi^2(1)$ statistics before taking their means. Effective sample size ( $N$ ) was estimated using the procedure outlined in the online supplement of Mallard et al. (2019).<sup>47</sup>

We identified only 2 genome-wide hits for the higher-order *p*-factor, both of which were in LD with univariate hits for MDD and SCZ (Table S21), and have been described in multiple external GWAS of psychiatric traits (Table S22). The *p*-factor was characterized by the highest level of heterogeneity by far, with 69 loci identified for Q_SNP_ (Table S23), 49 of which were in LD with hits on the four psychiatric factors from the correlated factors model. Despite few hits for *p*, its considerable mean *χ*^2^ (1.795) may be attributable to the aggregation of heterogenous signal across factors 1-4 in the hierarchical factor GWAS.

In summary, very few SNPs act on these 11 disorders in a manner consistent with the presence of a *p*-factor, whereas many SNPs act on the 11 disorders according to patterns that are significantly inconsistent with the presence of a *p*-factor. Moreover, the high average mean *χ*^2^ for the *p*-factor suggest that the paucity of factor hits is not attributable to low power. The observed pattern of SNP effects, in combination with the extensive heterogeneity in the pattern of correlations with biobehavioral traits reported earlier, suggests that a single, higher-order common factor of genetic risk for psychiatric disease has low plausibility and little pragmatic utility for understanding the shared genetic architecture of the disorders.

### Post-hoc Multivariate GWAS: Bifactor Specification of p

An alternative approach to modelling the *p*-factor is to specify a bifactor model.^6,7^ In the bifactor model, the *p*-factor and four domain-specific factors are specified to be orthogonal to one another and to directly predict the 11 disorders (Figure 1d). In contrast to the hierarchical model in which the relationship between *p* and the 11 disorders is mediated by the four lower-order factors, the bifactor model allows for direct associations between *p* and the 11 disorders. As the hierarchical model reflects a constrained version of the bifactor model, the bifactor model is always able to approximate the empirical genetic covariance as well as, or better than, the hierarchical model.^46^ Indeed, the bifactor model fit the data very well (*χ*^2^[28] = 120.35, AIC = 196.35, CFI = .982, SRMR = .062). Multivariate GWAS results using the bifactor model are presented here in order to more fully consider the utility of a *p*-factor, but are treated as exploratory and post-hoc.^46^

A multivariate GWAS with the bifactor *p*-factor as the GWAS target identified 66 independent hits, including the two hits for the hierarchical *p*-factor (Table S24). Among these 66 hits, 38 were in LD with hits from the correlated factors model, 8 hits were novel relative to univariate hits, and 7 hits were novel relative to both univariate or correlated factors hits. Three hits were novel for psychiatric traits more generally (Table S25). We identified 76 Q_SNP_ hits, 50 of which were in LD with hierarchical *p* Q_SNP_ hits (Table S26). Although the bifactor specification of *p* produced more factor hits than did the hierarchical specification, the pattern of results with respect to the large number of Q_SNP_ hits and high overall mean *χ*^2^ of Q_SNP_ was similar, and the LDSC genetic correlation across these two specifications of *p* was > .99. Collectively, these results indicate low utility of the *p*-factor for either the bifactor or hierarchical specification.

### Estimating Causal Effects of Problematic Alcohol Use on Psychiatric Disease Risk

One third of the Q_SNP_ discoveries from the correlated factors model appeared to operate through pathways unique to ALCH. This observation motivated an examination of the causal effects of ALCH on the disorders and factors using a form of multi-trait Mendelian randomization (MR) within the Genomic SEM framework. We ran two types of MR models: one using the Q_SNP_ variant in the ADH1B gene as a single instrumental variable for ALCH, and a second multi-variant MR approach using 8 loci identified from an independent ALCH discovery GWAS as instrumental variables.^48^ The multi-variant approach allowed for pleiotropic effects of the loci on additional disorders or factors where appropriate (Supplementary Results). Results from the ADH1B and multi-variant Genomic SEM-MR approaches tentatively supported a causal effect of ALCH on MDD and BIP (Supplementary Results; Figures S28-29). In these models, ALCH loadings on factors 2-4 were no longer significant, but the remaining disorders continued to load significantly on their respective factors. This indicates that although ALCH may have causal effects on risk for at least two different disorders, multiple causation by ALCH alone is not sufficient to fully account for the widespread patterning of statistical pleiotropy observed among the remaining disorders examined here.

## Discussion

We used genetic factor models to identify four broad factors (Neurodevelopmental, Compulsive, Psychotic, and Internalizing) that provide a reasonable model of the genetic correlations among 11 major psychiatric disorders, as estimated using the most recent GWAS summary data for individuals of European ancestry. We find that the Compulsive, Psychotic, and Internalizing factors are generally effective at describing the genetic relationship between psychiatric disorders at biobehavioral, functional genomic, and molecular levels of analysis.

Results were less consistent with the utility of a Neurodevelopmental disorders factor. For example, numerous biobehavioral traits differed in their genetic correlations with AUT to the point where its disorder-specific etiology must diverge substantially from those of the other disorders loading on this factor. The Neurodevelopmental disorders factor also exhibited much higher degrees of heterogeneity with respect to associations with individual SNPs, suggesting few variants conferring risk for these disorders are likely to operate through a more general factor.

Although the genetic correlations among the 11 disorders were somewhat consistent with the concept of a general *p*-factor, a hierarchical factor model that specified such a *p*-factor was found to offer limited biological insight, obscuring patterns of genetic correlations with external biobehavioral traits, the enrichment of pleiotropy within specific biological annotations, and the associations with individual variants. A bifactor model identified a larger number of GWAS hits for *p*, but similar to the hierarchical model exhibited a great deal of SNP-level heterogeneity. Given that a *p*-factor was found to be insufficient for accounting for patterns of multivariate associations the question arises: What processes gives rise to the moderate genetic correlations observed among the four, first-order factors? One possibility is that genetic correlations among the four factors arise from shared biology underlying pairwise combinations of factors, and not from any biology that is shared across all factors. Similarly, genetic correlations among the factors themselves may reflect pairwise combinations of shared biology among disorders that are not shared across all disorders within a factor.

In some circumstances, genetic correlations across disorders may arise from direct, potentially mutual, causation between the factor or disorder-specific liabilities and one another^49^ or reflect causation directly between the symptoms of different disorders.^50^ Based on significant locus-specific violations of the four factor model at loci relevant to ALCH, including a locus in the ADH1B gene, we incorporated Mendelian randomization into Genomic SEM models in order to estimate the direct causal effect of ALCH on risk for the other disorders. Both single- and multi-variant MR indicated causal effects of ALCH on MDD and BIP. The capability to combine MR and Genomic SEM in order to simultaneously model latent variables and direct effects between disorders vastly increases the scope of possible models that can be evaluated in future work.

In order to identify gene sets and categories in which pleiotropic risk variants for multiple disorders are disproportionally localized, we developed Stratified Genomic SEM, and applied it to 168 annotations, including 29 annotations representing protein-truncating variant (PTV)-intolerant (PI) genes, genes expressed in the human brain cells in the hippocampus and prefrontal cortex, and their intersection. We find that the intersection between PI genes and genes expressed in both excitatory and GABAergic neurons explained an outsized proportion of the genetic variance in the Psychotic disorders factor, which primarily indexes genetic covariance between SCZ and BIP. This offers critical insight into increasingly specific classes of genes relevant to shared risk across two disorders with high genetic overlap. Across the four correlated factors, we find that conserved regions are generally enriched. As enrichment in conserved annotations has been previously reported for both psychiatric traits and a host of other complex traits (e.g., cognitive function, anthropometric traits^41,43^), the current findings suggest that these annotations confer risk for individual disorders via highly pleiotropic variants relevant for many different domains of functioning.

It is important to note a number of limitations of the current analytic framework. Stratified Genomic SEM inherits the assumptions and limitations of traditional S-LDSC.^33^ This includes using an additive model of gene action that does not consider the role of epistatic effects, and only modelling the covariance among relatively common variant SNPs for which LD information is available. In future work, larger univariate GWAS coupled with Stratified Genomic SEM would allow for fitting qualitatively distinct structural models for individual annotations. It is conceivable that a simpler two-factor model may best describe genetic covariance in evolutionarily conserved regions, whereas a five-factor model may reflect the underlying architecture in genes that are intolerant to protein truncation. The statistical tools developed here allow us to test such hypotheses by relaxing the assumption that a single structural model characterizes the genetic relationships across psychiatric disorders.

We also note that the pattern of results reported here is likely to have been influenced by the composition of the GWAS cohorts included. Summary statistics from well powered GWASs spanning the wide range of psychiatric disorders investigated here were only consistently available for individuals of European ancestry. A major priority for continued work in this area will be increase the diversity of populations for which psychiatric GWAS are available. Recently developed methods for the stratified analysis of genetic correlations across ancestral populations will be invaluable for the analysis of such data.^51^

Moreover, our results may have been influenced by the phenotyping and case-ascertainment methods used methods used. For instance, we included data from have been influenced by the inclusion of GWAS cohorts relying primarily on self-report phenotypes,^28^ though sensitivity analyses suggested minimal differences when excluding GWAS that used self-report cohorts. Future analyses may benefit from evaluating these findings using a set of traits that is balanced with respect to statistical power. Future research may also benefit from further accounting for heterogeneity in how samples are ascertained and disorders are assessed.^52^ Application of detailed and standardized assessment protocols to large, representative samples would of course be ideal. More pragmatically, future work may apply multivariate genetic approaches, such as those showcased here, at the level of individual symptoms.^53^

The current analyses revealed four, correlated psychiatric factors that account for extensive genetic overlap across disorders. We evaluate and elucidate the composition of these factors by demonstrating patterns of correlations with external traits, develop and apply a novel method, Stratified Genomic SEM, to identify classes of genes that explain disproportionate levels of genetic covariance, and identify sets of loci with ranging levels of pleiotropy. We also estimate MR models where pleiotropy is a function of both latent factors and direct effects from one disorder liability on another. Our results offer critical insight into shared and disorder specific mechanisms of genetic risk and suggest possible avenues for revising a psychiatric nosology currently defined largely by clinical observation. Evidence derived from multivariate genetic analysis, alongside evidence at other levels of explanation (e.g., cognitive neuroscience, neurochemistry, environmental stressors), could guide future diagnostic revision.

## Method

### Overview of Genomic SEM and Stratified Genomic SEM

Genomic SEM is a two-stage Structural Equation Modelling approach. In the first stage, a genetic covariance matrix (*S*) and its associated sampling covariance matrix (*V*_*S*_) are estimated with a multivariate version of LD Score regression (LDSC). *S* consists of heritabilities on the diagonal and genetic covariances (co-heritabilities) on the off-diagonal. *V* consists of squared *SE*s of *S* on the diagonal and sampling covariances on the off-diagonal, which capture dependencies between estimating errors that will arise in situations such as participant sample overlap across GWAS phenotypes. In the second stage, a structural equation model is fit to *S* by optimizing a fit function that minimizes the discrepancy between the model-implied genetic covariance matrix (Σ(*θ*)) and *S*, weighted by the elements within *V*. We use the diagonally weighted least squares (WLS) fit function described in Grotzinger et al. (2019):^44^

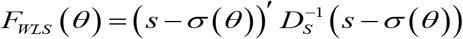

where *S* and Σ(*θ*) have been half-vectorized to produce *s* and σ(*θ*), respectively, and *D*_*S*_ is *V*_*S*_ with its off-diagonal elements set to 0. The sampling covariance matrix of the stage 2, Genomic SEM parameter estimates (*V*_*θ*_) are obtained using a sandwich correction described in Grotzinger et al. (2019):^44^

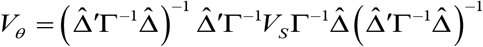

where 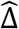 is the matrix of model derivatives evaluated at the parameter estimates, Γ is the stage 2 weight matrix, *D*_*S*_, and *V*_*S*_ is the sampling covariance matrix of *S*.

Stratified Genomic SEM extends this framework by allowing potentially different structural equation models to be fit to genetic covariance matrices estimated in different gene sets and categories. These gene sets and categories, collectively referred to as annotations, can be constructed based on a variety of sources, such as collateral gene expression data obtained from single-cell RNA sequencing. We develop a multivariate extension of Stratified LD Score Regression (S-LDSC)^33^ below to estimate these annotation-specific genetic covariance matrices and their associated sampling covariance matrices. We describe two types of annotation-specific genetic covariance matrices, *S*_*0*_ and *Sτ. S*_*0*_ contains estimates of genetic covariance within a specific annotation without controlling for overlap with other annotations. In other words, it is composed of the zero-order coefficients implied by the multivariate S-LDSC model. *Sτ* contains estimates of genetic covariance controlling for annotation overlap. In other words, it is composed of multiple regression coefficients estimated by the multivariate S-LDSC model. The distinction between *S*_*0*_ and *Sτ* directly parallels the distinction made in univariate S-LDSC^33^ between overall heritability explained by an annotation and the incremental contribution of a partition to heritability beyond all other annotations considered. Note that the estimates required to populate elements of an overall genome-wide *S* matrix can be produced either from the zero-order annotation that includes all SNPs or by aggregating parameters corresponding to each annotation from the multivariate S-LDSC model.

Below, we validate via simulation that Stratified Genomic SEM produces unbiased model parameter estimates and *SEs*, and that model fit indices appropriately favor the population generating model within a given annotation. There are a wide array of research questions that can be asked using Stratified Genomic SEM. In this paper, we examine genetic enrichment of variance in psychiatric genetic factors across a broad range of annotations.

### Multivariate Stratified LDSC

Under a multivariate extension of the S-LDSC model, the expected value of the product of *z* statistics for each pairwise combination of phenotypes for SNP *j* equals:

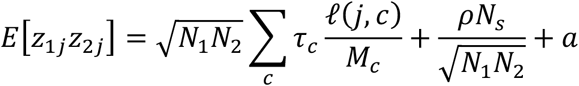

where *N*_*i*_ is the sample size for study *i, c* indexes a genomic annotation, *M*_*c*_ is the number of SNPs in annotation *c, ℓ*(*j,c*) is the LD score of SNP *j* with respect to annotation *c* (that is, the sum of squared LD this SNP has with all SNPs in the annotation), *τ*_*c*_ is a vector of free parameters used to compute the conditional contribution to heritability or coheritability (genetic covariance) in annotation *c, N*_*s*_ is the number of individuals included in both GWAS samples, *ρ* is the phenotypic correlation within the overlapping samples, and *a* is a term representing unmeasured sources of confounding such as shared population stratification across GWASs.^54^ The inclusion the term *M*_*c*_ in the above equation produces LD scores 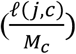 that are scaled relative to the size of the respective annotations, thereby allowing *τ*_*c*_ to be interpreted on the same scale as genome-wide estimates of heritability and coheritability, rather than on a per SNP scale. Note that when the *z* statistics for the same phenotype is double entered on the left hand side of the above equation, such that *E*[*z*_1*j*_*z*_2*j*_] becomes 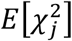, the equation reduces to the univariate S-LDSC model.^9^

Following Finucane et al. (2015),^33^ the multivariate S-LDSC model is estimated by regressing the product of *z* statistics against the annotation-specific LD scores using a weighted regression model (see online supplement of Finucane et al., 2015,^33^ for a description of how weights are calculated). Standard errors and dependencies among estimation errors (i.e., sampling covariances) are estimated using a multivariate block jackknife. As sample overlap creates a dependency between *z* statistics for the two traits, thus increasing their products, the S-LDSC intercept (*ρN*_*s*_*/*√*(N*_*1*_*N*_*2*_*) + a*) is affected, but the regression slope is unaffected, and the estimates of partitioned genetic covariance and their standard errors are not biased.

### Derivation of *S*_*τ*_ and *S*_*0*_

*S*_*τ,c*_ is a matrix containing estimates of genetic variance and covariance in annotation *c*, controlling for overlap with other annotations. It is composed of multiple regression coefficients, *τ*_*c*_, estimated directly with the multivariate S-LDSC model by populating each of its cells with the corresponding *τ* estimate from the multivariate S-LDSC model.

*S*_*0,c*_ is a matrix containing estimates of genetic covariance in annotation *c*, without controlling for overlap with other annotations. The elements *ζ*_*c*_ composing *S*_*0,c*_ can be derived from the *τ*_*c*_ estimates from the multivariate S-LDSC model in combination with knowledge of annotation overlap. Thus, the zero-order contribution of target annotation *t* to heritability or co-heritability is written as:

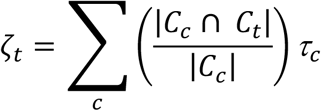

where |*C*_*c*_∩*C*_*t*_| is the number of SNPs in annotation *c* that are also in target annotation *t*, and |*C*_*c*_| is the total number of SNPs in annotation *c* (alternatively expressed as *M*_*c*_), such that 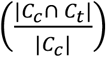 reflects the proportion of SNPs in annotation *c* that are also in target annotation *t*. This proportion is used to weight the term *τ*_*c*_ for each annotation in deriving the zero-order contribution of target annotation *t* to heritability or coheritability.

When the multivariate S-LDSC model is correct, *Sτ* is expected to produce unbiased estimates of the conditional contribution of an annotation to genetic covariance, after controlling for the effects of variants in all other annotation (i.e., accounting for the fact that variants can reside in multiple annotations). In comparison, *S*_*0*_ is expected to produce unbiased estimates of the total contribution of all genetic variants in an annotation to genetic covariance (i.e., irrespective of its overlap with the other annotations). *S*_*0*_ has two desirable properties. First, its estimate is not as directly contingent on which other annotations are included in the multivariate S-LDSC model. Second, because it does not decompose contributions of an annotation into those that are shared vs. unique of other annotations, it is expected to produce more stable estimates at small and moderate sample sizes. For this reason, the empirical Stratified Genomic SEM analyses reported here employ *S*_*0*_ matrices, and should be interpreted accordingly.

### Simulations of Stratified Genetic Covariance

#### Simulation Procedure

Using raw individual-level genotype data simulation, we sought to validate the point estimates and standard errors (*SE*s) produced by Stratified Genomic SEM. We compare results for *S*_*0*_ and *Sτ*. We began by generating 100 sets of 45, 100% heritable phenotypes (“orthogonal genotypes”) using the GCTA package.^55^ Each 100% heritable phenotype was specified to have 10,000 randomly selected causal variants from within a particular annotation. These phenotypes were paired with genotypic data for 100,000 randomly selected, unrelated individuals of European descent from UKB data for the 1,209,498 SNPs present in HapMap3.

The simulated genotypes were used to construct six different factor structures for six causal annotations. All orthogonal genotypes were scaled *M*=0, *SD*=1. For three of the causal annotations (DHS Peaks, H3K27ac, and PromoterUSC) seven genotypes for each annotation were used to construct six new correlated genotypes, each as the weighted linear combination of a domain-specific genetic factor and a general genetic factor, which was constructed from the seventh genotype. For the remaining three causal annotations (FetalDHS, H3K9ac, and TFBS) eight genotypes for each annotation were used to construct two sets of three correlated genotypes for two correlated general genetic factors, constructed from the seventh and eighth genotypes. A set of six “total” genotypes was created by summing a factor indicator genotype from each of the six causal annotations. As each genotype within each annotation was specified to have 10,000 causal SNPs, the “total’ genotypes created as the sum of six annotation had 60,000 causal SNPs in the population generating model.

Phenotypes were subsequently constructed as the weighted linear combination of one of the six “total” genotypes and domain-specific environmental factors (randomly sampled from a normal distribution with *M*=0, *SD*=1). Heritabilities for phenotypes 1-6 were all set to 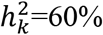, such that the weights for the genotypes were 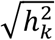 and the weights for the environmental factors were 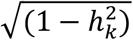. Each of the 600 phenotypes (100 sets of 6 phenotypes) was then analyzed as a univariate GWAS in PLINK^56^ to produce univariate GWAS summary statistics. The summary statistics were then munged, and Stratified Genomic SEM using the 1000 Genomes Phase 3 BaselineLD Version 2.2 model was used to construct 100 sets of 6×6 stratified zero-order genetic covariance matrices (*S*_*0*_), *τ* covariance matrices (*Sτ*), and their corresponding sampling covariance matrices (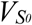 and *V*_S*τ*_).

#### Validating S_0_ and 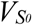

For the zero-order genetic covariance matrix, we would expect the annotation including all SNPs—i.e. the genome-wide annotation—to reflect the weighted linear combination of the generating covariance matrices specified for the six causal annotations, with weights equal to the proportion of all SNPs contained in each of the corresponding causal annotations. For each of the six causal annotations, we expect the zero-order covariance matrix for the corresponding annotation to be a linear combination of that annotation’s population-generating matrix and the remaining annotations’ population-generating matrices weighted by the proportion of SNPs overlapping across the annotations. To test these expectations, we created average observed covariance matrices across the 100 simulations for the genome-wide annotation and six causal annotations. The estimated *S*_*0*_ genome-wide covariance matrix approximately reflected an additive mixture of the six population generating covariance matrices, and was estimated with minimal bias (absolute value of mean discrepancy = .004; Figure S30b). In addition, the observed covariance matrices for each of the causal annotations were minimally biased relative to the generating population (Figure S30, Table S40).

In order to evaluate the accuracy of the *SEs*, we analyzed the ratio of the mean *SE* estimate across the 100 simulations over the empirical *SE* (calculated as the standard deviation of the parameter estimates across the 100 simulations). A value above 1 for this ratio indicates conservative *SE* estimates. This ratio was calculated within each of the annotations and for each cell of the covariance matrix. The average ratio across annotations and cells of the covariance matrix was 1.030 (Figure S31 for distribution across all annotations; Table S40 for ratio within causal annotations). Thus, we have produced a *SE* estimate for stratified heritability and covariance that performs as expected. In fact, our estimates are very slightly *conservative* as the mean *SE* was slightly larger than the empirical *SE*. Moreover, the average *z* statistic for heritability and covariance estimates within the causal annotations were all highly significant, suggesting more than adequate power under the conditions of the current simulation (Table S40).

#### Validating Sτ and V_Sτ_

The expectation for the genetic *S*_*τ*_ covariance matrices is that the observed covariance matrices will reflect the generating model within only that annotation. Indeed, the causal annotations closely matched their respective population generating covariance matrices and bias was minimal (Table S40; Figure S32). We then analyzed the ratio of the mean *SE* estimate across the 100 runs over the empirical *SE* (calculated as the standard deviation of the parameter estimates across the 100 runs). The average ratio of *SE* estimates was 1.014 across all annotations (Figure S31) and, importantly, was also close to 1 for the causal annotations (Table S40). Results for 4,459 of the total 5,300 covariance matrices produced negative heritability estimates. This included some of the causal annotations (Table S30), but was largely true for the non-causal annotations. Negative heritability estimates are unsurprising for the non-causal annotations as their population generating effect is 0. The *z* statistics for the *S*_*τ*_ heritabilities and covariances were, on average, smaller relative to the *S*_*0*_ covariance matrices (Table S40). This is to be expected as the *S*_*0*_ covariance matrices include power gained from variance shared with overlapping annotations.

The *S*_*τ*_ covariance matrices for the causal annotations were then used as input for Genomic SEM models. The two types of population generating models—a common factor and correlated factors model—were run for each annotation. For all causal annotations, Genomic SEM estimates closely matched the parameters specified in the generating population (Figure S33). In addition, the ratio of the mean model *SE*s over the empirical *SE*s was near 1. Model fit statistics (CFI, AIC, and model *χ*^2^) also generally favored the generating model for a particular annotation (Table S41). This was least true for the H3K27ac annotation. This is unsurprising as the population generating model for the H3K27ac annotation—a correlated factors model with a factor correlation of .7—most closely matched the competing common factor model. Collectively, these results indicate that stratified Genomic SEM produces unbiased parameter estimates and standard errors for *S*_*0*_ and *S*_*τ*,_ that *S*_*τ*_ shows specificity to the causal annotations of interest, and that model fit indices generally favor the appropriate model.

### Psychiatric Phenotypes

We curated the largest and most recent GWAS summary data from individuals of European ancestry for eleven major psychiatric disorders (Table S1). We refer the reader to the original articles for the corresponding univariate GWAS for details about sample ascertainment, quality control, and related procedures. For PTSD, MDD, ADHD, ANX, and ALCH, phenotype-specific meta-analyses of GWAS summary data derived from two different contributing sources per disorder were conducted in Genomic SEM so as to account for potentially unknown degrees of participant overlap across contributing samples. Models were specified to be equivalent to a fixed-effects meta-analysis, with both variables loading on the latent variable with an unstandardized loading fixed to 1.0, and both residual variances fixed to 0. LDSC-estimated genetic correlations within-phenotype-across-data-source were all ≥ .6 (Table S38). These GWAS meta-analyses in Genomic SEM were highly genetically correlated (≥ .94 as estimated with LDSC) with those estimated in METAL,^57^ which does not take sample overlap into account. Consistent with the differences in whether sample overlap is considered, Genomic SEM and METAL yielded univariate LDSC intercepts slightly below and slight above 1, respectively.

For the five meta-analyzed traits (Table S42) we provide Manhattan plots, tables of independent loci, and tables of hits that are in LD with hits previously identified in the GWAS catalogue (Figure S34; Table S43-S50). We find that many of the identified loci have been previously reported for the same or overlapping traits. As expected, the results for MDD and ADHD also overlap strongly with findings from the most recent MDD^58^ and ADHD^14^ papers that use highly similar samples to those that contributed summary data analyzed here. The observed differences are attributable to different analytic pipelines and partially non-overlapping contributing cohorts; for example, results reported from the published GWAS of ADHD^14^ include non-European samples, and hold some cohorts out for independent follow-up analyses.

While conducting this project the more recent PGC Freeze 2 release of PTSD became available.^59^ However, the GWAS *z* statistics and heritability estimates for PTSD Freeze 2 were lower than were observed for PTSD Freeze 1. As a result, our attempts to incorporate the PTSD Freeze 2 summary data produced a variety of technical problems (e.g. out of bounds genetic correlations and small heritability estimates). We therefore report results based on PTSD Freeze 1 summary data.

### Investigation of Genome-Wide Factor Structure

In order to explore the full-scope of factors solutions, EFAs were conducted using the *factanal* R package for two to five factor solutions using both oblique rotations, which allow for correlations among the latent factors, and orthogonal rotations, which assumes factors are independent (i.e., uncorrelated). Orthogonal rotations were examined as we, in part, sought to identify maximally separable dimensions with distinct sets of psychiatric indicators. EFAs were conducted for the genetic correlation structure derived from odd autosomes only. Confirmatory factor analyses (CFAs) specified on the basis of these EFAs were subsequently fit to a genetic correlation matrix estimated using only even autosomes. Using odd and even autosome covariance matrices for the exploratory and confirmatory models, respectively, provided a form of cross-validation to guard against model overfitting. For comparative purposes, we also consider model fit and final factor solutions for CFAs fit to the S-LDSC matrix (Figure S35).

For the CFAs, factors were assigned to traits when their standardized loading exceeded .35 in the corresponding EFAs, with two exceptions. First, for all EFAs with > 3 factors, a factor was identified with TS as its only indicator with standardized loading >.35. In the context of the CFAs, assigning TS to all factors at once, or to one factor at a time, resulted in issues with model convergence. Consequently, this final factor was removed in the CFA and TS was specified to always load on the factor with the largest EFA loading (excluding the factor defined only by TS) and models were compared where TS loaded onto one of the remaining factors. Among these combinations of TS models, a final model was selected using model fit indices (i.e., AIC, SRMR, and CFI). Second, for certain EFA solutions, there were traits that did not meet the standardized loading criteria of .35 for any factor. For these traits, we assigned factors to them in the CFA when their standardized loading exceeded a more lenient threshold of 0.2. We then inspected model fit indices for the follow-up CFA model to confirm that including those factor loadings provided better fit to the data.

All CFAs were fit using the Weighted Least Squares (WLS) estimator in the *GenomicSEM* R package. CFAs based on orthogonal EFA results allowed for freely correlated factors, as pruning factor loadings has the potential to reintroduce factor correlations. In the context of the CFAs, we also considered a common factor model in which all 11 traits loaded onto a single factor. CFAs with 4 correlated factors were similar in both factor structure and fit to the data (Table S51). In addition, the CFAs with 4 correlated factors provided far superior fit to the data (Figure S36), relative to the other models, with a number of the other CFAs failing to converge. Moreover, as indicated by model fit statistics, and observed directly in genetic correlation heatmaps, the correlation structure implied by the model estimates was much closer to the observed genetic correlations for these CFA solutions (Figure S37). The final model was chosen as a four correlated factor CFA (Table S52) as this ultimately provided the best fit to the data. Importantly, the model identified using a split of even and odd autosomes also fit the data well when applied to the genome-wide matrix estimated using autosomes 1-22 for LDSC (Figure 1b; *χ*^2^[33] = 161.66, AIC = 227.66, CFI = .975, SRMR = .072) and S-LDSC (Figure S35e; *χ*^2^[33] = 89.63, AIC = 155.63, CFI = .976, SRMR = .086).

The moderate factor correlations in this final model were also suggestive of a hierarchical structure (Figure S36d). This provided relatively comparable fit to the data for the LDSC genome-wide matrix (Figure 1c; *χ*^2^[35] = 171.37, AIC = 233.37, CFI = .974, SRMR = .079) and S-LDSC genome-wide matrix (Figure 35G; *χ*^2^[35] = 91.83, AIC = 153.83, CFI = .976, SRMR = .087). The absence of improved fit for the hierarchical model may reflect the fact that there was observable bias when comparing the factor correlations from the non-hierarchical model against the model implied correlations within the hierarchical model (Figure S37d).

### Genetic Correlations with Biobehavioral traits

For biobehavioral traits, summary statistics for 49 phenotypes broadly related to various domains of human health and well-being were downloaded from various online sources, primarily sourced from GWAS Atlas.^29^ For brain morphology, 101 summary statistics were downloaded from the GitHub page that corresponds to the summary data produced by Zhao et al. (2019).^30^ For accelerometer data, 24 summary statistics for each hour of movement across the day in UK Biobank were downloaded from the GCTA website.^31^ All summary statistics were cleaned and processed using the munge function of Genomic SEM, retaining all HapMap3 SNPs outside of the major histocompatibility complex (MHC) regions with minor allele frequencies (MAFs) ≥ .01. To evaluate potential associations between the psychiatric genetic factors and external traits, we used Genomic SEM to estimate genetic correlations between each of the four psychiatric factors, the hierarchical *p*-factor, and all of the relevant traits.

### Selection and Creation of Annotations

In order to construct the genome-wide S-LDSC matrix, and estimate stratified genetic covariance, we utilized pre-computed annotation files provided by the original S-LDSC authors.^33^ In line with recommendations, we utilized all annotations from the most recent 1000 Genomes Phase 3 BaselineLD Version 2.2^34^ that includes a total of 97 annotations ranging from coding, UTR, promoter, and flanking window annotations. For tissue specific histone marks, we included annotations constructed based on data from the Roadmap Epigenetics Project^35^ for narrowly defined peaks for DNase hypersensitivity, H3K27ac, H3K4me1, H3K4me3, H3K9ac, and H3K36me3 chromatin. For tissue specific gene expression, we include annotations constructed based on RNA sequencing data from human tissues from Genotype-Tissue Expression (GTEx)^36^ and for annotations constructed from human, mouse, and rat microarray experiments from the Franke Lab (i.e., DEPICT).^37^ For both tissue specific histone/chromatin marks and gene expression we utilized only brain and endocrine relevant regions in addition to 5 randomly selected control regions from each (i.e., 10 controls total).

We also created 29 annotations to examine the interaction between protein-truncating variant (PTV)-intolerant (PI) genes and human brain cells. PI genes were obtained from the Genome Aggregation Database (gnomAD), and ascertained using the probability of loss-of-function intolerance (pLI) metric. We selected genes with pLI > 0.9, producing a list of 3063 genes.^38^ Human brain cell gene sets were based on single-nucleus RNA-seq (sNuc-seq) data generated GTEx project brain tissues in the hippocampus and prefrontal cortex.^39^ Excluding sporadic genes and genes with low expression, for the 14 cell types we selected the top 1600 (∼15%) differentially expressed genes in each cell type, which likely cover all genes that are important for a specific cell type. PI × human brain cell gene sets contained the intersection of genes that are PTV-intolerant and each human brain cell gene set. Annotations were created using a 100kb window and LD information from the European subsample of 1000 Genomes Phase 3.

We do not estimate enrichment of psychiatric factors for continuous or flanking window annotations, yielding a total of 168 binary annotations across the baseline model, gene expression, histone marks, PI, and brain cell annotations. For a Bonferroni correction < .05 this corresponds to *p* < 2.98E-4. We note that continuous and flanking window annotations were retained for construction of the genome-wide, S-LDSC matrix.

### Estimating Genetic Enrichment of Model Parameters

We can examine whether the proportional contribution of an annotation to a given genome-wide parameter in Stratified Genomic SEM is different than would be expected on the basis of the relative size of that annotation, so long as the parameter is scaled comparably across all annotations considered.^60^ This is formalized by testing the null hypothesis,

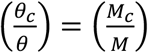

where *θ*_*c*_is the parameter estimate in annotation *c*, as estimated from a Genomic SEM model applied to *S*_*0,c*_; *θ* is the genome-wide parameter estimate, as estimated from a Genomic SEM model applied to the genome-wide *S* matrix derived via aggregating the conditional contributions of all annotations included in the multivariate S-LDSC model; *M*_*c*_ is the number of SNPs in annotation *c*; and *M* is the total number of SNPs. This formula can be rearranged to produce a ratio of ratios (the so-called *enrichment ratio*) that indexes the magnitude of enrichment:

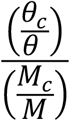

with a value of 1.0 corresponding to the null of no enrichment, values greater than 1.0 corresponding to enrichment (overrepresentation of signal in the annotation relative to its size), and values below 1.0 corresponding to depletion (underrepresentation of signal in the annotation relative to its size).

In the current application, we are interested in enrichment of pleiotropic and disorder-specific signal, as indexed by a factor model that allows the estimates of factor variances and disorder-specific uniquenesses, respectively, to vary across annotations, while holding all factor loadings invariant across annotations. We use a two-step model-fitting procedure to estimate the enrichment ratio in order to directly obtain an estimate of its *SE*. In Step 1, we estimate the factor loadings needed to scale the total genome-wide variances of the factors to 1.0. This is achieved by fitting a model to the genome-wide S-LDSC matrix in which unit variance identification is used. In Step 2, the loading estimates from the prior Step 1 model are fixed and the factor variance is freely estimated separately in each annotation using the *S*_*0,c*_ matrices. Thus, the estimated factor variances in Step 2 are scaled proportionally relative to the genome-wide factor variance (i.e., the numerator of the enrichment ratio). This estimate and its *SE* are subsequently divided by the proportion of SNPs in the corresponding annotation (i.e., the denominator of the enrichment ratio). For clarification, we note that genome-wide enrichment across all SNPs is exactly equal to 1. That is, for Step 2, if the genome-wide S-LDSC matrix is used as input, this produces a parameter estimate of 1, which is then divided by a proportion of 1.0, which reflects the ratio of *M/M* (i.e., all SNPs over all SNPs).

### Q_SNP_ Estimation

Q_SNP_ indexes violation of the null hypothesis that the SNP acts through a given factor. Put another way, it quantifies whether the individual SNP is more likely to operate through the common pathways of the psychiatric factors, or the independent pathways of individual disorders. The index thereby identifies loci that do not plausibly operate on the individual phenotypes exclusively by way of associations with common factor(s), and may be highly specific to the individual disorder. In the context of the multivariate GWAS for the correlated factors model, four separate follow-up models were estimated in which the SNP predicted three of the overarching factors *and* the indicators of the remaining fourth factor (see Figure S38 for path diagram). Comparing the model *χ*^2^ between the model in which the SNP predicted all four factors to one of these four, follow-up models produces a factor-specific Q_SNP._ For the hierarchical factor structure, we compared the model *χ*^2^ for a model in which the SNP predicted only the second-order *p*-factor, to the model *χ*^2^ for a model in which the SNP predicted only the four, first-order psychiatric factors. For the bifactor model, we compared a model in which the SNP predicted only the *p*-factor to a model in which in the SNP predicted both the *p*-factor and the remaining four orthogonal factors. For both the hierarchical and bifactor model, Q_SNP_ indexes heterogeneity at the level of the psychiatric factors (i.e., deviation from the null that the SNP operates through the *p*-factor). Therefore, a significant Q_SNP_ statistic for the hierarchical or bifactor model is likely to identify loci that are specific to a subset of the psychiatric factor(s). This is distinct from the interpretation of Q_SNP_ in the context of the correlated factors model as a significant hierarchical or bifactor Q_SNP_ may still conform to the local structure of one of the correlated factors.

### Quality Control Procedures

#### LD-Score Regression

Quality control (QC) procedures for producing the genetic covariance (*S*) and sampling covariance (*V*_*S*_) matrix followed the defaults in LDSC. This included removing SNPs with an MAF < 1%, information scores (INFO) < .9, SNPs from the MHC region, and filtering SNPs to HapMap3. The LD scores used for the analyses presented were estimated from the European sample of 1000 Genomes, but restricted to HapMap3 SNPs as these tend to be well-imputed and produce accurate estimates of heritability.

#### Multivariate GWAS

To obtain summary statistics for multivariate GWAS, we used the default QC procedures in Genomic SEM of removing SNPs with an MAF < .005 in the 1000 Genomes Phase 3 reference panel and SNPs with an INFO score < 0.6 in the univariate GWAS summary statistics. These are currently the default QC procedures for the *GenomicSEM* R package. Using these QC steps, there were 4,775,763 SNPs present across all eleven sets of European ancestry summary statistics. Prior to running any multivariate GWAS, all summary statistics were standardized with respect to the total variance in the outcome using the *sumstats* function in *GenomicSEM* and corrected for genomic inflation using the conservative approach of by multiplying of the standard errors by the univariate LDSC intercept when the intercept was above 1.

### Identification of Top Hits (Clumping) and Overlapping Hits

Lead SNPs for meta-analyzed univariate indicators and the latent genetic factors were identified using the clumping and pruning algorithm in FUMA.^61^ Independent significant SNPs were defined as crossing the genome-wide significance threshold of *p* < 5e-8 that were independent from other SNPs at *r*^*2*^ < 0.1. We used pre-calculated LD from European 1000 Genomes Phase 3 reference panel to identify independent SNPs. Top loci were subsequently identified by merging any SNPs in close proximity (< 250 kb) into a single genomic locus such that an individual locus could include multiple independent SNPs at *r*^*2*^ < 0.1. We depict only the significant loci (referred to as hits throughout the paper) in the Miami plots, but report independent significant SNPs in supplementary tables. This same pipeline was used for the full set of univariate summary statistics (i.e., not listwise deleted across all 11 traits) in order to produce a comparable set of loci for the univariate disorder GWAS. To determine overlap with hits across the factors and disorders, we identified all independent SNPs for the psychiatric factors that were in LD (*r*^*2*^ < 0.1) with independent SNPs for the individual disorders. As LD structure can vary across different cohorts, we also considered hits to be overlapping (in LD) if loci from the univariate disorder GWAS were within a 250 kb window (125 kb on either side of the index variant) of loci identified for the psychiatric factors or omnibus test.

## Data Availability

The data that support the findings of this study are all publicly available or can be requested for access. Specific download links for various datasets are directly below.

Summary statistics for data from the PGC can be downloaded or requested here: https://www.med.unc.edu/pgc/download-results/

Summary statistics for the Anxiety phenotype in UKB (TotANX_OR) can be downloaded here: https://drive.google.com/drive/folders/1fguHvz7l2G45sbMI9h_veQun4aXNTy1v

23andMe summary statistics are made available through 23andMe to qualified researchers under an agreement with 23andMe that protects the privacy of 23andMe participants. Please visit research.23andme.com/collaborate/#publication for more information

Summary statistics for the volume-based neuroimaging phenotypes were downloaded from: https://github.com/BIG-S2/GWAS

Summary statistics for the health and well-being complex trait correlations can be downloaded from: https://atlas.ctglab.nl/

Summary statistics for the circadian rhythm correlations across 24-hours can be downloaded from: https://cnsgenomics.com/software/gcta/#DataResource

Data from gnomAD used to identify PI genes for creation of annotations can be downloaded here: https://storage.googleapis.com/gnomad-public/release/2.1.1/constraint/gnomad.v2.1.1.lof_metrics.by_gene.txt.bgz

Gene count data per cell for creation of annotations were obtained from: https://storage.googleapis.com/gtex_additional_datasets/single_cell_data/GTEx_droncseq_hip_pcf.tar

Data which maps individual cells to cell types (e.g. neuron, astrocyte etc.) were obtained from: https://static-content.springer.com/esm/art%3A10.1038%2Fnmeth.4407/MediaObjects/41592_2017_BFnmeth4407_MOESM10_ESM.xlsx

Links to the LD-scores, reference panel data, and the code used to produce the current results can all be found at: https://github.com/MichelNivard/GenomicSEM/wiki

Links to the BaselineLD v2.2 annotations can be found here: https://data.broadinstitute.org/alkesgroup/LDSCORE/

## Code Availability

GenomicSEM software (which now includes the Stratified GenomicSEM extension), is an R package that is available from GitHub at the following URL: https://github.com/MichelNivard/GenomicSEM Directions for installing the GenomicSEM R package can be found at: https://github.com/MichelNivard/GenomicSEM/wiki

## Acknowledgements

This work presented here would not have been possible without the enormous efforts put forth by the investigators and participants from Psychiatric Genetics Consortium, iPSYCH, UK Biobank, and 23andMe. The work from these contributing groups was supported by numerous grants from governmental and charitable bodies as well as philanthropic donation. Research reported in this publication was supported by the National Institute Of Mental Health of the National Institutes of Health under Award Number R01MH120219. The content is solely the responsibility of the authors and does not necessarily represent the official views of the National Institutes of Health. ADG was additionally supported by NIH Grant R01HD083613. EMTD was additionally supported by NIH grants R01AG054628 and R01HD083613 and the Jacobs Foundation. EMTD is a faculty associate of the Population Research Center at the University of Texas, which is supported by NIH grant P2CHD042849. MGN is additionally supported by ZonMW grants 849200011 and 531003014 from The Netherlands Organisation for Health Research and Development, a VENI grant awarded by NWO (VI.Veni.191G.030) and is a Jacobs Foundation Fellow. WAA is supported by the “European Union’s Horizon 2020 research and innovation programme, Marie Sklodowska Curie Actions – MSCA-ITN-2016 – Innovative Training Networks under grant agreement No [721567]”. HFI is supported by the “Aggression in Children: unraveling gene-environment interplay to inform Treatment and InterventiON strategies” (ACTION) project. ACTION receives funding from the European Union Seventh Framework Program (FP7/2007-2013) under grant agreement no 602768. CML is supported by the National Institute for Health Research Biomedical Research Centre at South London and Maudsley NHS Foundation Trust and King’s College London. AMM is supported by the Wellcome Trust (104036/Z/14/Z, 216767/Z/19/Z), UKRI MRC (MC_PC_17209, MR/S035818/1). KPL is supported by the Deutsche Forschungsgemeinschaft (DFG: CRU 125, CRC TRR 58 A1/A5, No. 44541416), the European Union’s Seventh Framework Programme under Grant No. 602805 (Aggressotype), the Horizon 2020 Research and Innovation Programme under Grant No. 728018 (Eat2beNICE) and 643051 (MiND), Fritz Thyssen Foundation (No. 10.13.1185), ERA-Net NEURON/RESPOND, No. 01EW1602B, ERA-Net NEURON/DECODE, No. FKZ01EW1902 and 5-100 Russian Academic Excellence Project. GB is supported by the National Institute for Health Research Biomedical Research Centre at South London and Maudsley NHS Foundation Trust and King’s College London. PL is supported by NIH R01MH119243 and R00MH101367.

## Author Contributions

Study Design: Grotzinger, Nivard, Tucker-Drob

Methods Development: Grotzinger, Nivard, Tucker-Drob

Software Development: Grotzinger, Ip, Nivard, Tucker-Drob

Simulation Studies: Grotzinger, Nivard, Tucker-Drob

Gene Set and Annotation Creation: Akingbuwa, Grotzinger, Nivard

Genetic Factor Modelling, Multivariate GWAS, Complex Trait Correlations, and Multivariate

Enrichment Analyses: Grotzinger, Mallard, Nivard, Tucker-Drob

Writing: Grotzinger, Nivard, Tucker-Drob

Feedback and Editing: All authors contributed to editing the manuscript.

## Declaration of Interests

J.W.S. is an unpaid member of the Bipolar/Depression Research Community Advisory Panel of 23andMe. H.R.K. (Henry R. Kranzler) is a member of the American Society of Clinical Psychopharmacology’s Alcohol Clinical Trials Initiative, which was supported in the last three years by AbbVie, Alkermes, Ethypharm, Indivior, Lilly, Lundbeck, Otsuka, Pfizer, Arbor, and Amygdala Neurosciences. CML is on the SAB for Myriad Neuroscience. GB is a scientific advisor for COMPASS Pathways. The other authors declare no competing interests.

## Notes

### Author Declarations

We have used de-identified GWAS summary statistics for the analyses reported in the current study. For those no IRB oversight is necessary. Summary statistics used in the present analyses are largely publicly available for download using the database links noted in the manuscript. Summary statistics obtained for Major Depressive Disorder and ADHD were obtained from 23andMe after receiving approval for use in the present analyses. 23andMe research is IRB approved and all 23andMe Research participants provided consent for their data to be used in future analyses. A copy of the current 23andMe consent form can be found at: https://www.23andme.com/about/consent/.

